# Life Course Socioeconomic Position and health in older adulthood age: A Formal Mediation Analysis in the 1958 British Birth Cohort

**DOI:** 10.64898/2026.03.23.26349085

**Authors:** Yiling Guo, Alina Pelikh, George B Ploubidis, Alissa Goodman

## Abstract

**Background:** Childhood socioeconomic position (SEP) is a key determinant of later-life health. Understanding the extent to which adult SEP mediates this association into early old age is important for explaining how health inequalities are propagated across generations and how they might be addressed in later life. To our knowledge, no prospective study has examined whether childhood SEP remains associated with health at the threshold of older age and the extent to which any such association is mediated by adult SEP.

**Methods:** We used data from the 1958 British Birth Cohort, a prospective study that has followed participants since birth, drawing on earlier data collected at birth and ages 33 and 55 years and newly collected data from the age 62 sweep. Using interventional causal mediation analyses, we assessed whether adult occupational class, education, housing tenure, and income mediate associations between childhood social class (manual vs non-manual) and health at age 62 (self-rated health, C-reactive protein [CRP], cholesterol ratio, Glycated hemoglobin [HbA1c], and N-terminal pro–B-type natriuretic peptide [NT-proBNP]).

**Findings:** Associations between childhood SEP and self-rated health, CRP, cholesterol ratio, and HbA1c persisted after accounting for adult SEP. Mediation was outcome-specific and differed by sex. Among men, occupational class mediated 39% of the association with self-rated health (indirect effect RR 0·90, 95% CI 0·86–0·95) and education mediated 27% (0·93, 0·90–0·96). Among women, education mediated 10% (0·95, 0·91–0·98) and housing tenure mediated 6% (0·97, 0·94–0·99). Indirect effects for CRP were smaller, and mediation was minimal for cholesterol ratio, HbA1c, and NT-proBNP

**Interpretation:** Population-level improvements in adult SEP could reduce, but are unlikely to eliminate, later-life health inequalities associated with childhood SEP. Reducing these inequalities will require policies that address disadvantage in early life and improve adult financial and employment conditions.

**Funding:** UK Economic and Social Research Council

## Introduction

Population ageing presents a major challenge for health systems, pension and social policy. It is widely understood that health in older age is shaped by processes that begin early in life and unfold across the life course^1–2^, making it essential to understand the early-life determinants of healthy ageing. Evidence from British birth cohort shows a childhood SEP gradient in survival, with those from the disadvantaged childhood social class experiencing roughly 1.7-2.7 times higher premature mortality risk than those from more advantaged childhood social class^3^. Similar gradients have also been reported in other settings, such as the U.S. and Northern Europe^4^.

A central debate in life-course epidemiology concerns how much of this early-life imprint is amenable to intervention through adult socioeconomic circumstances, as one pathway among many. The critical period hypothesis posits that childhood SEP has independent effects on later health that are not fully modified by subsequent socioeconomic mobility^5^. In contrast, the accumulation and pathway hypotheses emphasize cumulative and sequential processes in which childhood SEP shapes adult SEP, which in turn influences later health^5^. In this framing, adult SEP represents one pathway linking childhood (dis)advantage to later health, with policy relevance for whether interventions should priorities early-life conditions, adult socioeconomic inequalities, or both?

Evidence suggests that childhood (dis)advantage influences later health through adult SEP and through effects that persist independent of adult SEP, suggesting that accumulation, pathway, and critical processes operate concurrently^6–8^. However, much of this evidence relies on traditional mediation approaches, inferring indirect effects either from attenuation of the exposure coefficients after adjustment (difference-in-coefficients approach)^7^, or from multiplying regression coefficients after adjustment (product-of-coefficients approach)^8^. These approaches require restrictive assumptions, including correctly specified linear models and no exposure-mediator interaction, and may yield biased estimates in complex life course settings characterized by non-linearity, interaction, and confounding^9^.

In the UK, much of the evidence to date on childhood SEP and health focuses on early or mid-adulthood, when socioeconomic trajectories and health risks are still unfolding^10–13^. Studies looking at health in older age have often relied on retrospectively reported childhood circumstances^14^, which do not prospectively trace socioeconomic trajectories across the full life course within the same individuals. To our knowledge, no prospective evidence addresses whether childhood SEP remains associated with health at the threshold of older age, and the extent to which any association is consistent with operating via adult SEP.

We aimed to fill these gaps by examining how the associations between childhood SEP and health at age 62 were mediated by adult SEP. Grounded in a counterfactual framework, we applied formal mediation methods to estimate interventional indirect effects via adult SEP and interventional direct effects representing the remaining association not operating through adult SEP as defined and measured in our study. Our estimates provide policy-relevant ‘what if’ contrasts by quantifying the potential reduction in later health inequalities under hypothetical shifts in adult SEP. Leveraging rich longitudinal data from the 1958 British Birth Cohort, this study provides rigorous empirical insight into the mechanisms linking childhood SEP to health at the onset of older age.

## Methods

### Data

The study used the National Child Development Study (NCDS, also known as the 1958 British birth cohort)^15–16^, a nationally representative longitudinal study of 17,415 individuals born in Britain during one week in 1958. To date, eleven sweeps of data are available, when cohort member were aged 0, 7, 11, 16, 23, 33, 42, 46, 50, 55, and 62 years. Information collected from age 0, 33, 55, 62 was used in this study. In 2020, 8,405 cohort members participated in the age 62 sweep, of whom 6,309 provided blood samples. The age 62 data collection took place over an extended period in 2020-2024, when study members were 62-66 years old.

### Outcome

We examined biomarkers reflecting systemic inflammation (C-reactive protein [CRP]) and cardiometabolic risk (cholesterol ratio, glycated haemoglobin [HbA1c], and N-terminal pro–B-type natriuretic peptide [NT-proBNP; a marker of heart failure]) in midlife, given their strong social patterning and established links to later morbidity and mortality^17–18^.

CRP (mg/L) values <3 mg/L were classified as no inflammation (0) and 3-10 mg/L as inflammation (1); values >10 mg/L were excluded to reduce acute infection effects. The cholesterol ratio was calculated as total cholesterol divided by high-density lipoprotein cholesterol (mmol/L). HbA1c was converted from mmol/mol to percentage units (%). NT-proBNP (pg/mL) was log-transformed and analysed as observed, as no physiologically justified correction exists, and medication effects are heterogeneous. We applied add-back corrections for medication use to approximate untreated cholesterol ratio and HbA1c levels among medicated participants (appendix section 4). For all biomarkers, higher values indicate poorer health. We also examined self-rated health, dichotomised as good (excellent/very good/good = 0) versus fair/poor (=1).

### Life course SEP

Childhood SEP was operationalised using parental occupational class at birth (Registrar General’s Social Class scheme, RGSC) and dichotomized as non-manual (I, II, and IIIN) versus manual (IIIM, IV, and V). Adult SEP at age 55 was operationalised using occupational class (manual/non-manual), housing tenure (owner/renter), educational attainment (degree or higher/no degree), and income to assess which distinct measures exert the strongest mediating effects.

### Directed acyclic graph and confounders

Figure 1 presents the directed acyclic graph (DAG) outlining the assumed causal relationships among confounders, childhood social class, adult SEP, and health outcomes. Childhood social class was the exposure, hypothesised to influence later-life health both directly and indirect via adult SEP. Separate analyses were conducted for each of the four adult SEP indicators specified as mediators.

**Figure. 1.**
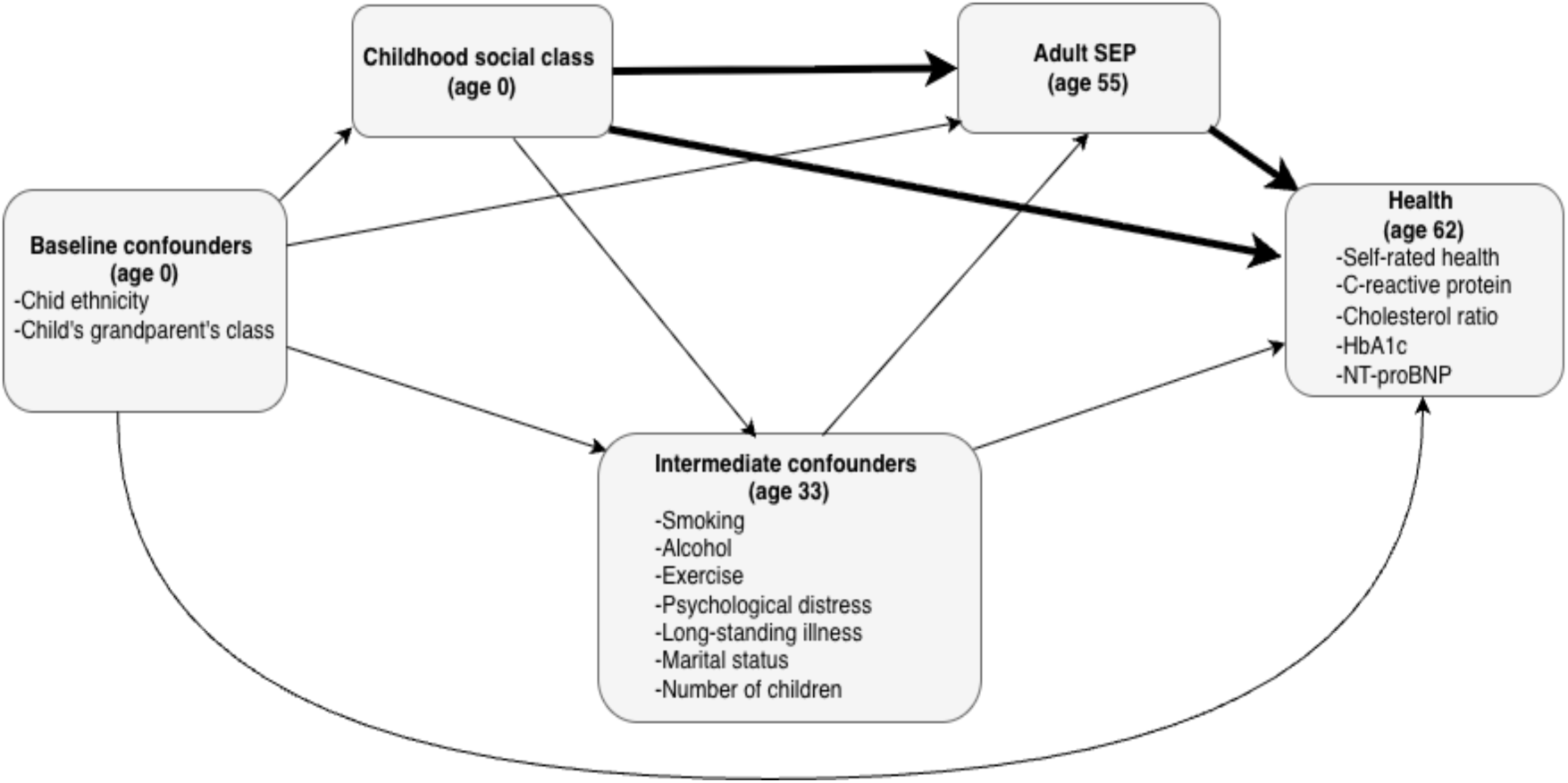
Directed acyclic graph (DAG) depicting the assumed causal structure, bold arrows indicate the estimands of interest.

Baseline confounders included ethnicity (Black and minority ethnic [BME]/non-BME), and grandparental social class (RGSC), reflecting demographic, and intergenerational social advantage that precede and may confound SEP trajectories and later health. Intermediate confounders, measured at age 33 and therefore prior to adult SEP at age 55, included smoking (yes/no), regular alcohol use (yes/no), low exercise (yes/no), psychological distress (standardized malaise score), long-standing illness (yes/no), marital status (married/not), and number of children. These variables may be affected by childhood SEP and can influence both adult SEP and later health. Sample characteristics are provided in the Appendix Table A1-A2.

Our primary estimands were the interventional indirect effect and direct effect. The interventional indirect effect captures the component operating via adult SEP. The interventional direct effect represents the remaining childhood SEP and health association not captured by adult SEP as measured here (and may reflect pathways operating through other mechanisms). Causal interpretation additionally assumes no unmeasured confounding conditional on variables included in Figure 1, as well as positivity, consistency, and no interference^19^.

### Statistical analysis

We first summarized age 62 health outcomes overall and by childhood social class. We then used regression models to examine the association between childhood social class and health outcomes, adjusting for baseline confounders. Additional models then assessed how childhood social class related to adult SEP and how adult SEP related to health outcomes, helping clarify the pathways linking early life social class to later health. We then conducted mediation analyses to estimate interventional direct and indirect effect of childhood social class on heath, adjusting for all confounders.

Interventional effects were estimated using a g-formula approach with direct counterfactual imputation. It allows identification of indirect effects despite exposure-induced intermediate confounding (i.e. midlife behavioural, health, and social factors in Figure 1), and without requiring the cross-world independence assumptions of natural effects^20–23^. It also yields estimates under well-defined hypothetical interventions, thereby clarifying the alternative scenarios being compared and enhancing the policy relevance of the analysis (Table 1).

**Table 1.** Interpreting patterns of interventional effects.

| Pattern of effects | Interpretation | Implications |
| --- | --- | --- |
| Small direct effect; large indirect effect | Most of the association operates via the mediator. | Improving adult SEP may substantially offset childhood disadvantage. |
| Large direct effect; small indirect effect | The association is unexplained by the mediator. | Intervening on adult SEP alone will be insufficient; early-life interventions are critical. |
| Both direct and indirect effect are moderate | Multiple pathways contribute meaningfully. | A combined early-life and adult-life strategy may be required. |
| Both direct and indirect effect are null | No clear mediated or direct pathway. | Interventions on either SEP unlikely to change the outcome. |

Mediation analyses were performed using the CMAverse R package^24^, with additional g-formula options provided by the CMAversePlus add-on^25^. Confidence intervals for all estimands were obtained via bootstrapping. All analyses were conducted separately for men and women using Stata 18.5 and R. For the regression-based analyses, missing data were imputed using multiple imputation by chained equations (MICE), generating 50 imputed datasets^26–28^. The mediation procedure implemented MICE internally (appendix section 2).

All analyses were restricted to participants with non-missing outcome data, and analytic sample sizes therefore varied across outcomes (appendix tables A1–A2). Regression and mediation analyses included 4183 men and 4180 women for self-rated health, 1996 men and 2018 women for C-reactive protein, 2479 men and 2505 women for cholesterol ratio, 2461 men and 2481 women for HbA1c, and 2413 men and 2473 women for NT-proBNP. Inverse probability weighting was applied to account for attrition. Models including adult occupational class as the mediator excluded participants who were out of the labour force at age 55 years, reducing the corresponding samples to 3715 men and 3347 women, 1787 men and 1649 women, 2220 men and 2034 women, 2204 men and 2015 women, and 2155 men and 2014 women, respectively.

## Results

Table 2 presents descriptive statistics based on participants with non-missing outcome data and observed childhood social class. Participants from manual childhood social class had a higher prevalence of fair or poor self-rated health than those from non-manual childhood social class. They also had a higher prevalence of inflammation, a less favorable mean cholesterol ratio, and higher mean HbA1c. Little difference was observed for log-transformed NT-proBNP. Similar patterns were seen in sex-stratified descriptive analyses (appendix table A3).

**Table 2.** Health outcomes by childhood social class.

|  | Overall | Non-manual | Manual | SEP difference (CI) |
| --- | --- | --- | --- | --- |
| **Outcome** |  |  |  |  |
| Poor/fair self-rated health, n (%) | 1933 (25.6) | 463 (19.3) | 1470 (28.6) | 9.3 (7.3, 11.3) |
| Presence of inflammation, n (%) | 803 (22.1) | 193 (16.1) | 610 (25.1) | 9.0 (6.3, 11.7) |
| Cholesterol ratio, mean (SD) | 4.03 (1.29) | 3.88 (1.22) | 4.11 (1.32) | 0.23 (0.15, 0.31) |
| Glycated hemoglobin (HbA1c) (%), mean (SD) | 6.05 (1.05) | 5.93 (0.82) | 6.11 (1.15) | 0.17 (0.11, 0.23) |
| NT-proBNP (pg/mL), mean (SD) | 4.15 (0.89) | 4.16 (0.88) | 4.15 (0.90) | -0.01 (-0.07, 0.04) |

Notes: Data are n (%) or mean (SD). SEP difference is manual minus non-manual and is expressed as an absolute percentage-point difference for binary outcomes and a mean difference for continuous outcomes. Denominators vary across outcomes because of outcome-specific missingness. NT-proBNP was log-transformed prior to analysis.

Table 3 presents regression estimates of the total association between childhood social class and health at age 62, adjusted for baseline confounders. Among men, a non-manual (vs manual) childhood social class was associated with 34% lower odds of poor self-rated health (OR = 0.66, 95% CI: 0.48, 0.91) and 46% lower odds of elevated C-reactive protein (OR = 0.54, 95% CI: 0.35,0.82). A non-manual childhood social class was also associated with more favorable cardiometabolic profiles, reflected in lower cholesterol ratio, glycated hemoglobin, and NT-proBNP levels. Among women, associations were in the same direction but generally stronger. Appendix Tables A6 and A7 show strong intergenerational socioeconomic persistence and graded associations between adult SEP and health at age 62, most clearly for self-rated health.

**Table 3.** Regression-based estimates of associations between childhood social class and health outcomes at age 62.

| **Panel A. Men** |  |  |  |  |  |
| --- | --- | --- | --- | --- | --- |
| | Self-rated health<br>OR (CI)<br>(n = 4,183) | C-reactive protein<br>OR (CI)<br>(n = 1,996) | Cholesterol ratio<br>$\beta$ (CI)<br>(n = 2,479) | Glycated haemoglobin<br>$\beta$ (CI)<br>(n = 2,461) | NT-proBNP<br>$\beta$ (CI)<br>(n = 2,413) |
| Childhood social class<br>(ref: manual) | 0.66 (0.48, 0.91) | 0.54 (0.35, 0.82) | -0.11 (-0.33, 0.11) | -0.16 (-0.37, 0.05) | 0.95 (0.81, 1.13) |
| Grandparental class<br>(ref: manual) | 0.74 (0.52, 1.06) | 0.93 (0.58, 1.49) | 0.04 (-0.20, 0.28) | -0.08 (-0.29, 0.14) | 0.85 (0.71, 1.01) |
| BME (ref: not BME) | 1.37 (0.66, 2.84) | 0.32 (0.09, 1.08) | 0.44 (-0.70, 1.58) | 0.96 (-0.79, 2.72) | 0.76 (0.50, 1.17) |
| Intercept | 0.56 (0.48, 0.65) | 0.32 (0.26, 0.40) | 4.33 (4.23, 4.44) | 6.24 (6.13, 6.34) | 4.06 (3.97, 4.15) |
| **Panel B. Women** |  |  |  |  |  |

| | Self-rated health<br>OR (CI)<br>(n = 4,180) | C-reactive protein<br>OR (CI)<br>(n = 2,018) | Cholesterol ratio<br>$\beta$ (CI)<br>(n = 2,505) | Glycated haemoglobin<br>$\beta$ (CI)<br>(n = 2,481) | NT-proBNP<br>$\beta$(CI)<br>(n = 2,473) |
| --- | --- | --- | --- | --- | --- |
| Childhood social class<br>(ref: manual) | 0.49 (0.37, 0.65) | 0.51 (0.37, 0.70) | -0.35 (-0.48, -0.21) | -0.09 (-0.19, -0.00) | 1.01 (0.91, 1.13) |
| Grandparental class<br>(ref: manual) | 0.74 (0.55, 1.01) | 0.96 (0.67, 1.37) | -0.09 (-0.25, 0.07) | -0.22 (-0.31, -0.13) | 0.94 (0.84, 1.06) |
| BME (ref: not BME) | 0.97 (0.44, 2.16) | 0.88 (0.31, 2.45) | -0.14 (-0.50, 0.22) | 0.03 (-0.16, 0.22) | 0.76 (0.61, 0.95) |
| Intercept | 0.63 (0.55, 0.73) | 0.42 (0.36, 0.50) | 3.91 (3.83, 3.99) | 6.05 (5.98, 6.13) | 4.37 (4.31, 4.43) |

Notes: BME = Black and minority ethnic. CI = 95% confidence interval. NT-proBNP was log-transformed prior to analysis; exponentiated coefficients represent ratios of geometric means. Models are adjusted for baseline confounders.

The effect of childhood social class on health outcomes remained strong after mediation analysis was applied. Among men (Table 4), for self-rated health, the interventional direct effects (RR ≈ 0.78–0.87 across mediators) indicated that holding men’s adult class to the distribution it would take under disadvantaged childhood class, shifting childhood class from disadvantaged to advantaged reduced fair/poor self-rated health by 13-22%. The interventional indirect effects implied that if everyone were born into disadvantaged class, shifting adult class to the distribution observed among those from advantaged childhood class would reduce the risk of fair/poor self-rated health by approximately 10% (RR = 0.90, 95% CI: 0.86, 0.95; 39% mediated). Smaller indirect effects were observed via education (RR = 0.93, 95% CI: 0.90, 0.96; 27% mediated), housing (RR = 0.97, 95% CI: 0.94, 0.99; 11% mediated), and income (RR = 0.99, 95%CI: 0.97, 1.00; 4% mediated).

**Table 4.** Interventional direct and indirect effects of childhood social class on health outcomes at age 62, men.

| Mediator | Direct effect | Indirect effect | Overall effect | rPM |
| --- | --- | --- | --- | --- |
| **Self-rated health RR (CI)** |  |  |  |  |
| Class | 0.87 (0.73, 1.00) | 0.90 (0.86, 0.95) | 0.79 (0.67, 0.91) | 0.39 (0.15, 1.10) |
| Housing | 0.79 (0.69, 0.90) | 0.97 (0.94, 0.99) | 0.77 (0.67, 0.87) | 0.11 (0.03, 0.29) |
| Education | 0.83 (0.73, 0.93) | 0.93 (0.90, 0.96) | 0.77 (0.67, 0.87) | 0.27 (0.10, 0.52) |
| Income | 0.78 (0.68, 0.88) | 0.99 (0.97, 1.00) | 0.77 (0.67, 0.88) | 0.04 (0.00, 0.11) |
| **C-reactive protein RR (CI)** |  |  |  |  |
| Class | 0.65 (0.50, 0.86) | 0.91 (0.83, 0.99) | 0.59 (0.46, 0.76) | 0.14 (0.01, 0.50) |
| Housing | 0.63 (0.49, 0.85) | 1.00 (0.96, 1.03) | 0.62 (0.49, 0.84) | — |
| Education | 0.68 (0.54, 0.93) | 0.91 (0.86, 0.96) | 0.62 (0.49, 0.84) | 0.16 (0.07, 0.43) |
| Income | 0.62 (0.49, 0.85) | 1.00 (0.99, 1.01) | 0.62 (0.49, 0.84) | — |
| **Cholesterol ratio $\beta$ (CI)** |  |  |  |  |
| Class | -0.15 (-0.30, 0.02) | 0.00 (-0.05, 0.05) | -0.15 (-0.27, 0.01) | — |
| Housing | -0.12 (-0.27, 0.02) | -0.01 (-0.04, 0.00) | -0.14 (-0.28, 0.01) | — |
| Education | -0.10 (-0.24, 0.05) | -0.04 (-0.07, -0.01) | -0.14 (-0.28, 0.01) | — |
| Income | -0.14 (-0.28, 0.01) | 0.00 (-0.01, 0.01) | -0.14 (-0.28, 0.01) | — |
| **Glycated hemoglobin $\beta$ (CI)** |  |  |  |  |
| Class | -0.10 (-0.22, 0.03) | -0.02 (-0.06, 0.01) | -0.12 (-0.22, -0.01) | — |
| Housing | -0.14 (-0.24, -0.02) | -0.01 (-0.03, 0.01) | -0.14 (-0.24, -0.03) | — |
| Education | -0.14 (-0.24, -0.02) | -0.01 (-0.03, 0.02) | -0.14 (-0.24, -0.03) | — |
| Income | -0.14 (-0.24, -0.04) | 0.00 (-0.01, 0.01) | -0.14 (-0.24, -0.03) | — |
| **NT-proBNP estimates $\beta$ (CI)** |  |  |  |  |
| Class | 0.03 (-0.07, 0.12) | -0.02 (-0.05, 0.01) | 0.01 (-0.09, 0.10) | — |
| Housing | 0.04 (-0.03, 0.14) | 0.00 (-0.01, 0.02) | 0.04 (-0.03, 0.14) | — |
| Education | 0.04 (-0.04, 0.14) | 0.00 (-0.02, 0.03) | 0.04 (-0.03, 0.14) | — |
| Income | 0.04 (-0.03, 0.14) | 0.00 (-0.01, 0.00) | 0.04 (-0.03, 0.14) | — |

Notes: RR = risk ratio. CI = 95% confidence interval. rPM = randomised analogue of proportion mediated. Overall effect and rPM are reported for completeness. rPM is reported only when the overall effect is meaningfully different from zero and the interventional direct and indirect effects operate in the same direction with a non-negligible indirect effect.

A similar pattern was observed for C-reactive protein. Direct effects remained substantial (RR ≈ 0.63–0.68 across mediators), indicating a strong remaining childhood social class gradient in elevated CRP after accounting for adult SEP among men. Indirect effects were evident via occupational class and education (RR ≈ 0.91; 14–16% mediated), implying that shifting these adult SEP from the disadvantaged to the advantaged childhood distributions would reduce inflammation risk by around 9%. For cholesterol ratio, glycated hemoglobin (HbA1c), and NT-proBNP, interventional indirect effects were close to the null, suggesting that these childhood social class gradients were largely not captured by adult SEP measure here.

Among women (Table 5), direct effects for self-rated health were also sizable (RR ≈ 0.66–0.69 across mediators), indicating that holding women’s adult class to the distribution it would take under disadvantaged childhood class, shifting childhood class from disadvantaged to advantaged reduced fair/poor self-rated health by 31–34%. Indirect effects were smaller than among men and were most evident via education (RR = 0.95, 95% CI: 0.91, 0.98; 10% mediated), with limited mediation via housing (6% mediated) and income (3% mediated), and little evidence via occupational class. For C-reactive protein, direct effects again remained sizeable (RR ≈ 0.62–0.69), with limited mediation, mostly evidently via housing (RR = 0.97; 6% mediated). Indirect effects were small for cholesterol and HbA1c.

**Table 5.** Interventional direct and indirect effects of childhood social class on health outcomes at age 62, women.

| Mediator | Direct effect | Indirect effect | Overall effect | rPM |
| --- | --- | --- | --- | --- |
| **Self-rated health RR (CI)** |  |  |  |  |
| Class | 0.68 (0.59, 0.80) | 0.97 (0.93, 1.01) | 0.66 (0.57, 0.78) | 0.05 ( -0.01, 0.16) |
| Housing | 0.67 (0.59, 0.77) | 0.97 (0.94, 0.99) | 0.65 (0.56, 0.75) | 0.06 (0.02, 0.12) |
| Education | 0.69 (0.59, 0.79) | 0.95 (0.91, 0.98) | 0.65 (0.56, 0.75) | 0.10 (0.03, 0.20) |
| Income | 0.66 (0.57, 0.76) | 0.98 (0.96, 1.00) | 0.65 (0.56, 0.75) | 0.03 (0.01, 0.08) |
| **C-reactive protein RR (CI)** |  |  |  |  |
| Class | 0.62 (0.47, 0.77) | 1.04 (1.00, 1.09) | 0.65 (0.50, 0.80) | — |
| Housing | 0.68 (0.56, 0.81) | 0.97 (0.92, 1.00) | 0.66 (0.55, 0.79) | 0.06 (0.00, 0.18) |
| Education | 0.69 (0.56, 0.82) | 0.96 (0.92, 1.01) | 0.66 (0.55, 0.79) | 0.07 (-0.03, 0.21) |
| Income | 0.66 (0.54, 0.78) | 1.02 (0.99, 1.05) | 0.67 (0.56, 0.79) | — |
| **Cholesterol ratio $\beta$ (CI)** |  |  |  |  |
| Class | -0.28 (-0.38, -0.14) | 0.00 (-0.03, 0.01) | -0.28 (-0.37, -0.16) | — |
| Housing | -0.28 (-0.38, -0.15) | -0.01 (-0.03, 0.00) | -0.29 (-0.39, -0.16) | — |
| Education | -0.29 (-0.39, -0.17) | 0.00 (-0.02, 0.02) | -0.29 (-0.39, -0.17) | — |
| Income | -0.29 (-0.39, -0.17) | 0.00 (-0.01, 0.01) | -0.29 (-0.39, -0.16) | — |
| **Glycated hemoglobin $\beta$ (CI)** |  |  |  |  |
| Class | -0.15 (-0.22, -0.06) | 0.00 (-0.02, 0.01) | -0.16 (-0.23, -0.07) | — |
| Housing | -0.13 (-0.21, -0.06) | -0.01 (-0.02, 0.00) | -0.13 (-0.21, -0.07) | — |
| Education | -0.13 (-0.22, -0.06) | 0.00 (-0.02, 0.01) | -0.13 (-0.22, -0.07) | — |
| Income | -0.13 (-0.21, -0.07) | 0.00 (-0.01, 0.00) | -0.13 (-0.21, -0.07) | — |
| **NT-proBNP $\beta$ (CI)** |  |  |  |  |
| Class | 0.04 (-0.05, 0.13) | -0.01 (-0.03, 0.00) | 0.03 (-0.06, 0.11) | — |
| Housing | 0.02 (-0.06, 0.08) | 0.00 (-0.01, 0.01) | 0.02 (-0.05, 0.08) | — |
| Education | 0.03 (-0.05, 0.09) | -0.01 (-0.02, 0.01) | 0.02 (-0.05, 0.08) | — |
| Income | 0.02 (-0.06, 0.09) | 0.00 (-0.01, 0.01) | 0.02 (-0.05, 0.08) | — |

Notes: RR = risk ratio. CI = 95% confidence interval. rPM = randomised analogue of proportion mediated. Overall effect and rPM are reported for completeness. rPM is reported only when the overall effect is meaningfully different from zero and the interventional direct and indirect effects operate in the same direction with a non-negligible indirect effect.

To examine the robustness of our main findings, we completed several post-hoc analyses. First, we tested alternative outcome specifications: C-reactive protein as a continuous outcome (appendix table A10), cholesterol ratio as a binary outcome (table A11), and HbA1c as a binary outcome (table A12). Second, we examined alternative medication assumptions and restrictions, including alternative statin adjustments for total cholesterol (table A13) and exclusion of participants taking heart-failure-related medications in NT-proBNP analyses (table A14). Third, we imposed sample restrictions, including restriction to a consistent 2022–23 biomarker collection window (tables A15–A16) and restriction of self-rated health analyses to blood-sample providers (table A17). Fourth, we assessed whether income mediation findings were sensitive to alternative operationalization of income, including binary income measures based on different thresholds (tables A18–A19). Fifth, we examined the sensitivity of income mediation findings to possible measurement error by using income measured at a different timepoint (tables A20–A21). Main results were substantively unchanged in these analyses.

## Discussion

Three central findings emerged from our analyses. First, childhood SEP showed strong independent influence on self-rated health, inflammatory marker (CRP), and cardiometabolic biomarkers (cholesterol ratio and HbA1c) more than six decades later. Second, mediation through adult SEP was modest and outcome-specific: adult SEP accounted for some share of the association for self-rated health and inflammatory marker, but little of the association for cardiometabolic biomarkers. Third, mediation differed by adult SEP indicators and sex: among men, occupational class and education explained a sizable share of the association, whereas among women the overall association was stronger but less explained by the adult SEP measures included.

The strong associations between childhood SEP and health were reflected in sizable interventional direct effects. Even under a hypothetical equalisation of adult SEP, a substantial childhood SEP on health gradient remained. This patten was evident across subjective (self-rated health) and objective (CRP, cholesterol ratio, and HbA1c) health measures, strengthening the interpretation that early-life (dis)advantage has strong independent biological and health consequences that are independent of mid-life adult SEP. These findings align with developmental origins and life-course epidemiological frameworks, which emphasize early-life material and psychosocial conditions as foundational determinants of later-life health^1–2^.

Mediation effect was outcome specific. For self-rated health, indirect effects were evident, particularly through occupational class and education among men and through education and housing tenure among women. There was also some evidence of mediation for C-reactive protein, similarly through education among men and housing among women. For cholesterol ratio, HbA1c, and NT-proBNP, indirect effects were close to null. We think that these results should not be interpreted as evidence that adult SEP has no effect on later-life cardiometabolic health. Rather, they suggest that, under the specific hypothetical shifts in adult SEP considered here, the childhood SEP gradient in these biomarkers would change only modestly, pointing to mechanisms not captured by adult SEP alone.

Our findings are consistent with a broader life-course literature documenting that both childhood and adult SEP matter for health in mid-life^6–8^. Earlier NCDS analyses at age 44/45 have reported independent and comparably sized associations of childhood and adult social class with midlife cardiometabolic and inflammatory biomarkers in mutually adjusted models^10,13^. Extending follow up to older age, we found that adult SEP accounts for only a small share of the childhood gradient in later-life cardiometabolic outcomes, suggesting a more modest mediating role at the threshold of older age than might be inferred from midlife evidence. This conclusion was consistent in simpler mutually adjusted models, where controlling for adult SEP only modestly reduced childhood SEP associations (results available upon request), suggesting that the limited mediation is not an artefact of the interventional approach but reflects the pattern in the data at this life-course stage.

Two potential explanations may account for the weaker role of adult SEP in our study. First, by age 62 cardiometabolic risk may increasingly reflect the long-term biological embedding of childhood disadvantage. Early-life adversity may contribute to cumulative physiological wear and tear that becomes more apparent with age and is not easily offset by later socioeconomic circumstances. Thus, even if adult SEP remains relevant, it may account for a smaller share of the childhood gradient in early older age than has been observed in mid-life. Second, selective survival may also have attenuated the observed mediating role of adult SEP. By age 62, the analytical sample is likely to be more selected, as individuals with the most disadvantaged socioeconomic trajectories and poorest health are less likely to survive. This may compress socioeconomic variation among those observed at older ages and reduce the extent to which adult SEP appears to account for childhood differences in cardiometabolic health.

The contrasting pattern observed across outcomes is informative. A larger share of the childhood SEP gradient in self-rated health was captured by adult SEP. Among objective biomarkers, mediation patterns varied: we observed evidence of mediation for CRP, whereas mediation was limited for cardiometabolic markers. One plausible explanation is that self-rated health is a broad evaluative construct encompassing psychological and mental wellbeing alongside physical functioning^17^ and that the dimensions people draw on when rating their health may vary across social groups^29^. This may make self-rated health more responsive to adult socioeconomic position measured close to outcome assessment. In contrast, cardiometabolic markers reflect longer-term physiological embedding of early life conditions^30^. Consistent with this interpretation, prior work on mental health has similarly suggested mediation via adult SEP^31^, including evidence from the same cohort used here^32^. Future research should test whether similar patterns emerge across other health outcomes and physiological systems.

The mediation patterns also differed by adult SEP indicators and sex. Among men, occupational class and education explained a sizable share of the association between childhood SEP and self-rated health and C-reactive protein, suggesting that labor-market position and educational attainment were important pathways linking early-life SEP to later health. Among women, mediation was weaker overall. Occupational class explained little of the association, while education and housing tenure accounted for only a modest share, leaving most of the association between childhood SEP and health not captured by the adult SEP measures considered. The somewhat greater contribution of housing tenure than women’s occupational class suggests that material living conditions and household-level advantage may have been more relevant pathways than individual labor-market position for women in this cohort.

In the 1958 cohort, occupational class among men often reflected continuous full-time employment and thus serve as a reasonable proxy for cumulative adult socioeconomic exposure. For women, however, occupational trajectories were often characterized by part-time work, labor-force interruptions, and weaker attachment to the labor market^33^. Women’s occupational class may therefore have captured their adult material and social circumstances less completely. By contrast, housing tenure, as a household-level indicator, may have better reflected the economic context in which many women in this generation lived, particularly where resources were pooled within couples or mediated through partners.

Given the limited mediation observed for several outcomes, it is likely that pathways beyond the adult SEP measures considered here contribute to the childhood SEP gradient in later-life health. Fundamental cause theory posits that socioeconomic advantage operates through access to flexible resources, such as knowledge, power, material capacity, and social connections, that enable individuals to deploy varied strategies to protect health as risks and opportunities evolve over time^34^. From this perspective, the substantial direct effects in our analyses are best interpreted as the remaining childhood SEP and health association not captured by the adult SEP indicator examined, rather than implying an absence of underlying mechanisms. More broadly, early-life socioeconomic conditions pattern cumulative environmental exposures, socially structured stress processes, access to supportive social networks, and interactions with institutions, which may contribute to the remaining association observed here.

Several limitations should be acknowledged. Adult SEP was measured at a single midlife time point, which may understate cumulative socioeconomic exposure across adulthood. Additionally, although the interventional mediation framework improves causal interpretability relative to conventional approaches, it relies on the assumption that all relevant confounders of the exposure–outcome, exposure–mediator, and mediator–outcome relationships have been adequately captured. Given the long temporal span between childhood SEP, adult SEP, and later-life health, some residual confounding from unmeasured early-life or long-term psychosocial and environmental processes can be expected.

Notwithstanding these limitations, this study makes methodological and substantive contributions to life-course epidemiology and ageing research. Drawing on over six decades of prospective data from the 1958 British Birth Cohort, we examined the long-term direct and indirect effects of childhood SEP on health at age 62 operating through adult SEP, capturing inequalities across an unusually long temporal horizon. Through a counterfactual interventional mediation framework, we further moved beyond conventional approaches that rely on strong and often implausible assumptions regarding exposure–mediator independence. This approach allows us to estimate policy-relevant decompositions of the childhood SEP and health association while accommodating exposure-induced mediator–outcome confounding^35^.

Taken together, our findings suggest that, if policies were able to shift adult SEP at the population level in the manner represented by our hypothetical contrasts, some reduction in later-life health inequalities would be expected, although such shifts are unlikely to fully eliminate the health gradient associated with childhood SEP. In practice, the most realistic levers for shifting adult SEP at scale are population-level policies affecting financial and employment conditions, alongside early-life interventions to reduce disadvantage. The implication of these ‘what if’ contrasts depend on the extent to which comparable shifts in adult SEP are feasible within the prevailing social and policy context^36^.

## Data Availability

All original datasets from the 1958 NCDS are freely available to download at the UK Data Archive https://ukdataservice.ac.uk/.

## Contributors

All authors defined the research question and designed the study. Y.G. performed the statistical analyses and wrote the first draft. All authors critically contributed to the interpretation of the results, revised the manuscript, and approved the final version for publication.

## Data sharing

All original datasets from the 1958 NCDS are freely available to download at the UK Data Archive.

## Acknowledgements

The Economic and Social Research Council funds the Centre for Longitudinal Studies (CLS) Resource Centre (ES/W013142/1) which provides core support for the CLS cohort studies. While the CLS Resource Centre makes these data available, CLS does not bear any responsibility for the analysis or interpretation of these data by researchers. The CLS cohorts are only possible due to the commitment and enthusiasm of their participants, their time and contribution are gratefully acknowledged. AG, GBP, and AP were supported by the UCL Centre for Longitudinal Studies [ES/W013142/1], GBP was also supported by the ESRC Centre for Society and Mental Health at King’s College London [ES/S012567/1]. The views expressed are those of the authors and not necessarily those of the ESRC, UCL or King’s College London

## Supplementary material

### 1. Descriptive statistics

**Table A1.** Self-rated health sample characteristics based on complete cases.

| Variable | Men |  | Women |  |
| --- | --- | --- | --- | --- |
|  | N | Mean or % | N | Mean or % |
| Self-rated health at 62 |  |  |  |  |
| Good | 3,095 | 74.0 | 3,098 | 74.1 |
| Fair/poor | 1,088 | 26.0 | 1,082 | 25.9 |
| Total | 4,183 |  | 4,180 |  |
| Childhood social class |  |  |  |  |
| Manual | 2,579 | 67.7 | 2,586 | 68.6 |
| Non-manual | 1,230 | 32.3 | 1,183 | 31.4 |
| Total | 3,809 |  | 3,769 |  |
| Social class at 55 |  |  |  |  |
| Manual | 1,164 | 37.5 | 670 | 23.3 |
| Non-manual | 1,939 | 62.5 | 2,203 | 76.7 |
| Total | 3,103 |  | 2,873 |  |
| Economic activity at 55 |  |  |  |  |
| Inactive | 471 | 13.0 | 839 | 22.0 |
| Active | 3,166 | 87.0 | 2,971 | 78.0 |
| Total | 3,637 |  | 3,810 |  |
| Housing tenure at 55 |  |  |  |  |
| Renter | 397 | 19.3 | 458 | 19.0 |
| Owner | 1,657 | 80.7 | 1,953 | 81.0 |
| Total | 2,054 |  | 2,411 |  |
| Education at 55 |  |  |  |  |
| Non-degree qualifications | 2,727 | 76.2 | 2,881 | 76.7 |
| Degree or higher university degree | 850 | 23.8 | 876 | 23.3 |
| Total | 3,577 |  | 3,757 |  |
| Income at 55 (log transformed) | 2,802 | 10.1 | 2,618 | 10.0 |
| Child's ethnicity |  |  |  |  |
| Not BME | 4,107 | 97.8 | 4,112 | 97.8 |
| BME | 92 | 2.2 | 94 | 2.2 |
| Total | 4,199 |  | 4,206 |  |
| Grandparental class |  |  |  |  |
| Manual | 2,359 | 71.8 | 2,371 | 71.7 |
| Non-manual | 927 | 28.2 | 936 | 28.3 |
| Total | 3,286 |  | 3,307 |  |
| Smoking |  |  |  |  |
| No | 2,514 | 71.3 | 2,654 | 71.2 |
| Yes | 1,011 | 28.7 | 1,072 | 28.8 |
| Total | 3,525 |  | 3,726 |  |
| Regular alcohol use |  |  |  |  |
| No | 970 | 27.5 | 1,928 | 51.7 |
| Yes | 2,563 | 72.5 | 1,803 | 48.3 |
| Total | 3,533 |  | 3,731 |  |
| Low exercise |  |  |  |  |
| No | 2,807 | 79.7 | 2,946 | 79.3 |
| Yes | 717 | 20.3 | 770 | 20.7 |
| Total | 3,524 |  | 3,716 |  |
| Having long-standing illness or not |  |  |  |  |
| No | 2,943 | 83.4 | 3,225 | 86.5 |
| Yes | 585 | 16.6 | 502 | 13.5 |
| Total | 3,528 |  | 3,727 |  |
| Marital status |  |  |  |  |
| Married | 2,435 | 71.1 | 2,700 | 73.8 |
| Not married | 989 | 28.9 | 961 | 26.2 |
| Total | 3,424 |  | 3,661 |  |
| Malaise score (standardized) | 3,528 | 0.2 | 3,723 | -0.1 |
| Number of child | 3,201 | 1.4 | 3,495 | 1.6 |
| N in Main Survey | 4,199 |  | 4,206 |  |

Notes: Analytic sample sizes for self-rated health: men n = 4,183; women n = 4,180. Models treating adult social class as the mediator excluded participants who reported being out of the labour force at age 55, resulting in reduced analytic samples: men n = 3,715; women n = 3,347

**Table A2.** Biomarker sample characteristics based on complete cases.

| Variable | Men |  | Women |  |
| --- | --- | --- | --- | --- |
|  | N | Mean or % | N | Mean or % |
| C-reactive protein (CRP) at 62 |  |  |  |  |
| Presence of inflammation | 374 | 18.7 | 512 | 25.4 |
| No inflammation | 1,622 | 81.3 | 1,506 | 74.6 |
| Total | 1,996 |  | 2,018 |  |
| HDL cholesterol (mmol/L) at 62 | 2,479 | 1.4 | 2,507 | 1.7 |
| Total cholesterol (mmol/L) at 62 | 2,484 | 5.7 | 2,506 | 6.1 |
| Cholesterol ratio at 62 | 2,479 | 4.3 | 2,505 | 3.8 |
| *Glycated hemoglobin (HbA1c) (%)* at 62 | 2,461 | 6.1 | 2,481 | 6.0 |
| NT-proBNP (pg/mL) at 62 (log transformed) | 2,413 | 4.0 | 2,473 | 4.3 |
| Childhood social class |  |  |  |  |
| Manual | 1,930 | 67.3 | 1,925 | 67.8 |
| Non-manual | 937 | 32.7 | 913 | 32.2 |
| Total | 2,867 |  | 32.2 |  |
| Social class at 55 |  |  |  |  |
| Manual | 886 | 36.3 | 527 | 23.1 |
| Non-manual | 1,556 | 63.7 | 1,759 | 76.9 |
| Total | 2,442 |  | 2,286 |  |
| Economic activity at 55 |  |  |  |  |
| Inactive | 354 | 12.5 | 610 | 20.5 |
| Active | 2,484 | 87.5 | 2,361 | 79.5 |
| Total | 2,838 |  | 2,971 |  |
| Housing tenure at 55 |  |  |  |  |
| Renter | 289 | 18.4 | 322 | 17.4 |
| Owner | 1,282 | 81.6 | 1,527 | 82.6 |
| Total | 1,571 |  | 1,849 |  |
| Education at 55 |  |  |  |  |
| Non-degree qualifications | 2,122 | 75.1 | 2,241 | 75.8 |
| Degree or higher university degree | 702 | 24.9 | 714 | 24.2 |
| Total | 2,824 |  | 2,955 |  |
| Income at 55 (log transformed) | 2,250 | 10.1 | 2,100 | 10.1 |
| Child's ethnicity |  |  |  |  |
| Not BME | 3,089 | 98.1 | 3,088 | 97.7 |
| BME | 59 | 1.9 | 73 | 2.3 |
| Total | 3,148 |  | 3,161 |  |
| Grandparental class |  |  |  |  |
| Manual | 1,777 | 71.5 | 1,772 | 71.3 |
| Non-manual | 707 | 28.5 | 712 | 28.7 |
| Total | 2,484 |  | 2,484 |  |
| Smoking |  |  |  |  |
| No | 2,011 | 73.3 | 2,094 | 72.8 |
| Yes | 731 | 26.7 | 782 | 27.2 |
| Total | 2,742 |  | 2,876 |  |
| Regular alcohol use |  |  |  |  |
| No | 772 | 28.1 | 1,472 | 51.1 |
| Yes | 1,977 | 71.9 | 1,408 | 48.9 |
| Total | 2,749 |  | 2,880 |  |
| Low exercise |  |  |  |  |
| No | 2,208 | 80.6 | 2,276 | 79.3 |
| Yes | 533 | 19.4 | 594 | 20.7 |
| Total | 2,741 |  | 2,870 |  |
| Having long-standing illness or not |  |  |  |  |
| No | 2,298 | 83.7 | 2,489 | 86.5 |
| Yes | 447 | 16.3 | 389 | 38.9 |
| Total | 2,745 |  | 2,878 |  |
| Marital status |  |  |  |  |
| Married | 1,924 | 72.2 | 2,093 | 74.1 |
| Not married | 742 | 27.8 | 732 | 25.9 |
| Total | 2,666 |  | 2,825 |  |
| Malaise score (standardized) | 2,746 | 0.2 | 2,877 | -0.0 |
| Number of child | 2,487 | 1.4 | 2,696 | 1.6 |
| N in Main Survey | 3,148 |  | 3,161 |  |

Notes: Analytic sample sizes were C-reactive protein: men n = 1,996; women n = 2,018. cholesterol ratio: men n = 2,479; women n = 2,505). glycated haemoglobin: men n = 2,461; women n = 2,481. NT-proBNP: men n = 2,413; women n = 2,473. Models treating adult social class as the mediator excluded participants who reported being out of the labour force at age 55, resulting in reduced analytic samples: C-reactive protein (men n = 1,787; women n = 1,649); cholesterol ratio (men n = 2,220; women n = 2,034); glycated haemoglobin (men n = 2,204; women n = 2,015); NT-proBNP (men n = 2,155; women n = 2,014).

**Table A3.**
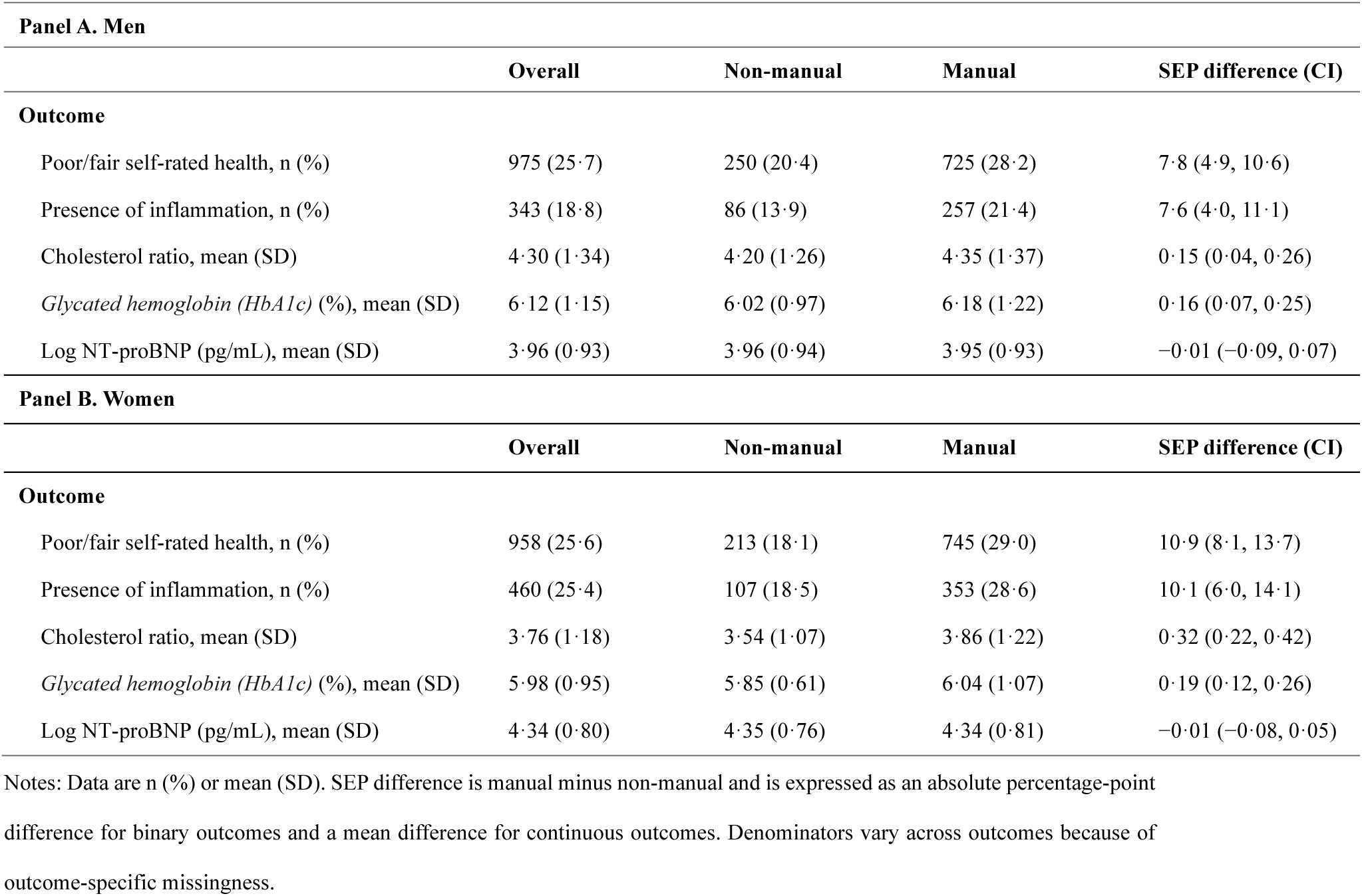
Age 62 health outcomes by childhood social class among men and women.

| **Panel A. Men** |  |  |  |  |
| --- | --- | --- | --- | --- |
|  | **Overall** | **Non-manual** | **Manual** | **SEP difference (CI)** |
| **Outcome** |  |  |  |  |
| Poor/fair self-rated health, n (%) | 975 (25.7) | 250 (20.4) | 725 (28.2) | 7.8 (4.9, 10.6) |
| Presence of inflammation, n (%) | 343 (18.8) | 86 (13.9) | 257 (21.4) | 7.6 (4.0, 11.1) |
| Cholesterol ratio, mean (SD) | 4.30 (1.34) | 4.20 (1.26) | 4.35 (1.37) | 0.15 (0.04, 0.26) |
| *Glycated hemoglobin (HbA1c) (%)*, mean (SD) | 6.12 (1.15) | 6.02 (0.97) | 6.18 (1.22) | 0.16 (0.07, 0.25) |
| Log NT-proBNP (pg/mL), mean (SD) | 3.96 (0.93) | 3.96 (0.94) | 3.95 (0.93) | −0.01 (−0.09, 0.07) |
| **Panel B. Women** |  |  |  |  |
|  | **Overall** | **Non-manual** | **Manual** | **SEP difference (CI)** |
| **Outcome** |  |  |  |  |
| Poor/fair self-rated health, n (%) | 958 (25.6) | 213 (18.1) | 745 (29.0) | 10.9 (8.1, 13.7) |
| Presence of inflammation, n (%) | 460 (25.4) | 107 (18.5) | 353 (28.6) | 10.1 (6.0, 14.1) |
| Cholesterol ratio, mean (SD) | 3.76 (1.18) | 3.54 (1.07) | 3.86 (1.22) | 0.32 (0.22, 0.42) |
| *Glycated hemoglobin (HbA1c) (%)*, mean (SD) | 5.98 (0.95) | 5.85 (0.61) | 6.04 (1.07) | 0.19 (0.12, 0.26) |
| Log NT-proBNP (pg/mL), mean (SD) | 4.34 (0.80) | 4.35 (0.76) | 4.34 (0.81) | −0.01 (−0.08, 0.05) |

Notes: Data are n (%) or mean (SD). SEP difference is manual minus non-manual and is expressed as an absolute percentage-point difference for binary outcomes and a mean difference for continuous outcomes. Denominators vary across outcomes because of outcome-specific missingness.

### 2. Missing data strategy

Missing data due to non-response are unavoidable in longitudinal studies and can reduce sample size, statistical power, and representativeness. Under Rubin’s framework, missingness may be missing completely at random (MCAR), missing at random (MAR), or missing not at random (MNAR) (Little & Rubin, 1989, 2002). Previous work using the NCDS has shown that non-response is systematically related to observed characteristics (Hawkes & Plewis, 2006; Mostafa et al., 2020), indicating that missingness is unlikely to be MCAR. As in most longitudinal cohort studies, missingness in the NCDS is therefore plausibly MAR or MNAR.

For the regression-based analyses, we performed Multiple Imputation with chained equations with 50 imputed datasets. We perform two multiple imputations: the first one on all the individuals who took part in the age 62 main survey (n = 8,405) to run the regressions on self-rated health; the second one on all the individuals who took part in the biomedical survey (n = 6,309) to run the regressions on biomarker outcomes. We then performed our regression analysis only on those with non-missing outcome (n = 8,363 for self-rated health; n = 4,014 for CRP; n =4,984 for cholesterol ratio; n = 4,942 for HbA1c; n = 4,886 for NT-proBNP), as suggested by Von Hippel (2007). Table A4-A5 report the proportion imputed for each variable used in the analysis.

The imputation model included all variables in the substantive models, as well as auxiliary variables. These auxiliary variables are variables associated with the outcomes and the probability of taking part in the survey, which include mother’s age at age 0, number of persons per room at age 0, father’s education, social problems (alcoholism etc.) at age 7, ever breastfed at age 7, cognitive ability summary at age 7, cognitive ability summary at age 11, conduct problems at age 11, child receiving help sch-backwardness at age 16, child’s school attendance at age 16, child drank alcohol at age 16, mathematics comprehension test score at age 16, conduct problems at age 16, voted for in 1979 general election at age 23, telephone in home at age 33, amount of physical effort in job at age 33, voted for 1987 General Election at age 33, housing tenure at age 33, social capital score at age 33, whether participated in NCDS V at age 42, membership in organizations at age 42 employer provided Pension or not at age 50

**Table A4.** The proportion of missing information in each variable in the self-rated health sample (N = 8,405)

| Variable | N missing | % missing |
| --- | --- | --- |
| Self-rated health | 42 | 0.50 |
| Adult social class | 2429 | 28.90 |
| Childhood social class | 827 | 9.84 |
| Child's sex | 0 | 0 |
| Child's ethnicity | 0 | 0 |
| Grandparental class | 1812 | 21.56 |
| Smoking | 1154 | 13.73 |
| Regular alcohol use | 1141 | 13.58 |
| Low exercise | 1165 | 13.86 |
| Malaise score | 1154 | 13.73 |
| Long-standing illness | 1150 | 13.68 |
| Marital status | 1320 | 15.70 |
| Number of children | 1709 | 20.33 |

**Table A5.** The proportion of missing information in each variable in the biomarker sample (N = 6,309)

| Variable | N missing | % missing |
| --- | --- | --- |
| C-reactive protein (CRP) | 2295 | 36.38 |
| HDL cholesterol | 1323 | 36.38 |
| Total cholesterol | 1319 | 20.91 |
| Cholesterol ratio | 1325 | 21.00 |
| *Glycated hemoglobin (HbA1c)* | 1367 | 21.67 |
| NT-proBNP | 1423 | 22.56 |
| Childhood class | 604 | 9.57 |
| Adult class | 1581 | 25.06 |
| Child's sex | 0 | 0 |
| Child's ethnicity | 0 | 0 |
| Grandparental class | 1341 | 21.26 |
| Smoking | 691 | 10.95 |
| Regular alcohol use | 680 | 10.78 |
| Low exercise | 698 | 11.06 |
| Malaise score | 686 | 10.87 |
| Long-standing illness | 686 | 10.87 |
| Marital status | 818 | 12.97 |
| Number of children | 1126 | 17.85 |

### 3. Association between study variables

**Table A6.** Regression-based estimates of associations between childhood social class and adult socioeconomic position.

| **Panel A. Men** |  |  |  |  |
| --- | --- | --- | --- | --- |
|  | **Adult social class** | **Housing tenure** | **Education** | **Income** |
|  | **OR (CI)** | **OR (CI)** | **OR (CI)** | **β (CI)** |
| Self-rated health analytic sample |  |  |  |  |
| Childhood social class (ref: manual) | 2.82 (2.09, 3.81) *** | 1.38 (0.91, 2.10) | 3.01 (2.19, 4.12) *** | 0.23 (0.01, 0.45) * |
| C-reactive protein analytic sample |  |  |  |  |
| Childhood social class (ref: manual) | 2.35 (1.58, 3.49) *** | 1.46 (0.85, 2.52) | 2.35 (1.65, 3.35) *** | 0.07 (-0.16, 0.30) |
| Cholesterol ratio analytic sample |  |  |  |  |
| Childhood social class (ref: manual) | 2.45 (1.71, 3.50) *** | 2.50 (1.80, 3.48) *** | 2.50 (1.80, 3.48) *** | 0.10 (-0.12, 0.31) |
| Glycated haemoglobin analytic sample |  |  |  |  |
| Childhood social class (ref: manual) | 2.52 (1.75, 3.61) *** | 1.52 (0.92, 2.51) | 2.51 (1.80, 3.51) *** | 0.10 (-0.11, 0.31) |
| NT-proBNP analytic sample |  |  |  |  |
| Childhood social class (ref: manual) | 2.49 (1.73, 3.57) *** | 1.50 (0.91, 2.48) | 2.58 (1.84, 3.60) *** | 0.11 (-0.10, 0.33) |
| **Panel A. Women** |  |  |  |  |
| Self-rated health analytic sample |  |  |  |  |
| Childhood social class (ref: manual) | 2.39 (1.66, 3.42) *** | 1.73 (1.20, 2.49) ** | 2.72 (2.07, 3.58) *** | 0.24 (0.04, 0.43) * |
| C-reactive protein analytic sample |  |  |  |  |
| Childhood social class (ref: manual) | 1.98 (1.25, 3.13) ** | 1.48 (0.88, 2.51) | 2.62 (1.85, 3.70) *** | 0.15 (-0.06, 0.37) |
| Cholesterol ratio analytic sample |  |  |  |  |
| Childhood social class (ref: manual) | 1.94 (1.24, 3.04) ** | 1.69 (1.05, 2.72) * | 2.51 (1.82, 3.47) *** | 0.17 (-0.02, 0.37) |
| Glycated haemoglobin analytic sample |  |  |  |  |
| Childhood social class (ref: manual) | 1.96 (1.25, 3.07) ** | 1.60 (0.99, 2.58) | 2.49 (1.80, 3.44) *** | 0.17 (-0.04, 0.37) |
| NT-proBNP analytic sample |  |  |  |  |
| Childhood social class (ref: manual) | 1.94 (1.24, 3.05) ** | 1.65 (1.02, 2.65) * | 2.52 (1.82, 3.49) *** | 0.16 (-0.04, 0.37) |

Notes: CI = 95% confidence interval. \*p < .05. \*\*p < 0.01. \*\*\*p < .001. Income was log-transformed (ln[income + 1]) prior to analysis. Models are adjusted for baseline confounders.

**Table A7.**
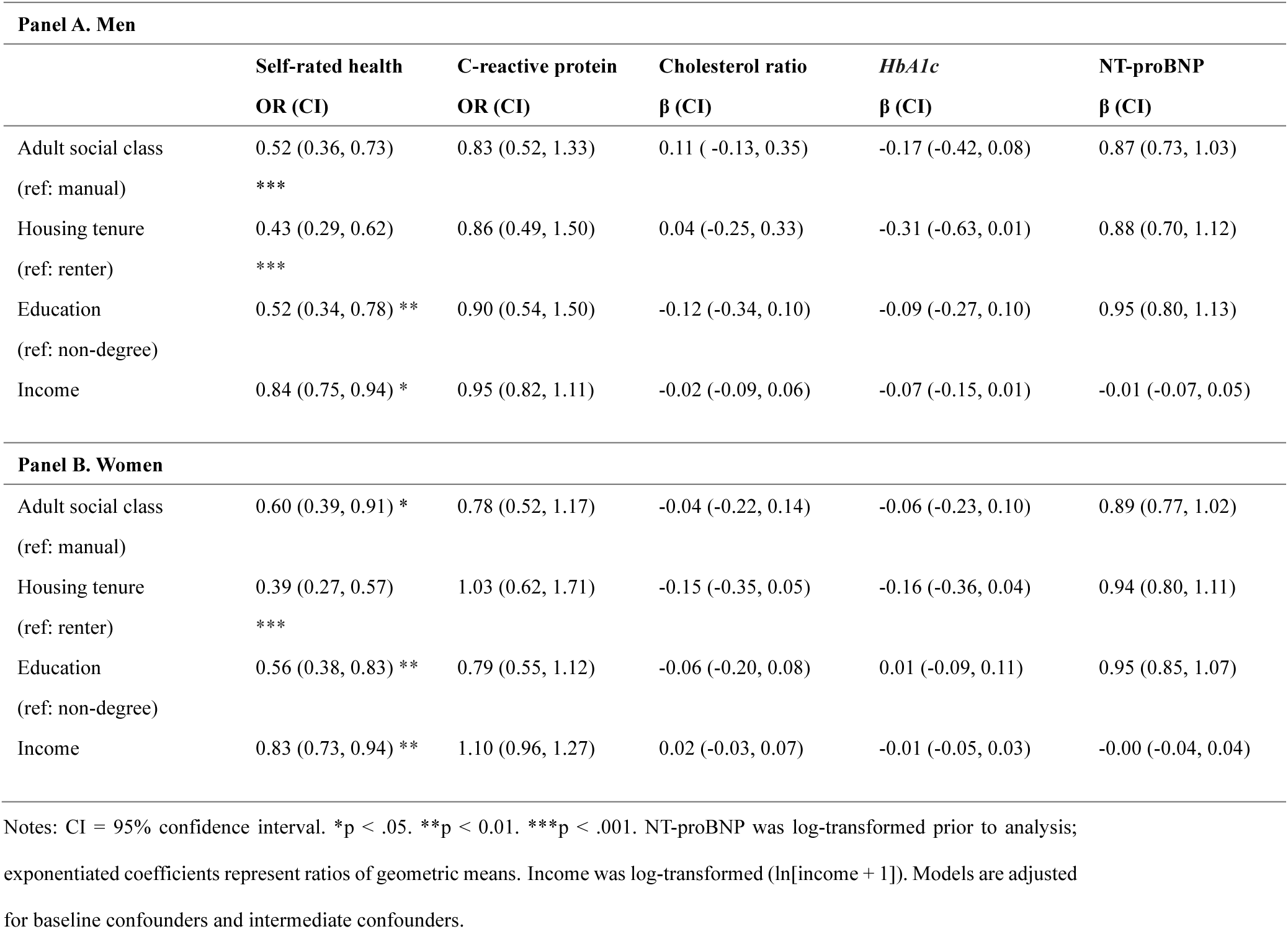
Regression-based estimates of associations between adult socioeconomic position and health outcomes at age 62.

| **Panel A. Men** |  |  |  |  |  |
| --- | --- | --- | --- | --- | --- |
|  | **Self-rated health** | **C-reactive protein** | **Cholesterol ratio** | **HbA1c** | **NT-proBNP** |
|  | **OR (CI)** | **OR (CI)** | **β (CI)** | **β (CI)** | **β (CI)** |
| Adult social class<br>(ref: manual) | 0.52 (0.36, 0.73) *** | 0.83 (0.52, 1.33) | 0.11 ( -0.13, 0.35) | -0.17 (-0.42, 0.08) | 0.87 (0.73, 1.03) |
| Housing tenure<br>(ref: renter) | 0.43 (0.29, 0.62) *** | 0.86 (0.49, 1.50) | 0.04 (-0.25, 0.33) | -0.31 (-0.63, 0.01) | 0.88 (0.70, 1.12) |
| Education<br>(ref: non-degree) | 0.52 (0.34, 0.78) ** | 0.90 (0.54, 1.50) | -0.12 (-0.34, 0.10) | -0.09 (-0.27, 0.10) | 0.95 (0.80, 1.13) |
| Income | 0.84 (0.75, 0.94) * | 0.95 (0.82, 1.11) | -0.02 (-0.09, 0.06) | -0.07 (-0.15, 0.01) | -0.01 (-0.07, 0.05) |
| **Panel B. Women** |  |  |  |  |  |
| Adult social class<br>(ref: manual) | 0.60 (0.39, 0.91) * | 0.78 (0.52, 1.17) | -0.04 (-0.22, 0.14) | -0.06 (-0.23, 0.10) | 0.89 (0.77, 1.02) |
| Housing tenure<br>(ref: renter) | 0.39 (0.27, 0.57) *** | 1.03 (0.62, 1.71) | -0.15 (-0.35, 0.05) | -0.16 (-0.36, 0.04) | 0.94 (0.80, 1.11) |
| Education<br>(ref: non-degree) | 0.56 (0.38, 0.83) ** | 0.79 (0.55, 1.12) | -0.06 (-0.20, 0.08) | 0.01 (-0.09, 0.11) | 0.95 (0.85, 1.07) |
| Income | 0.83 (0.73, 0.94) ** | 1.10 (0.96, 1.27) | 0.02 (-0.03, 0.07) | -0.01 (-0.05, 0.03) | -0.00 (-0.04, 0.04) |

Notes: CI = 95% confidence interval. \*p < .05. \*\*p < 0.01. \*\*\*p < .001. NT-proBNP was log-transformed prior to analysis; exponentiated coefficients represent ratios of geometric means. Income was log-transformed (ln[income + 1]). Models are adjusted for baseline confounders and intermediate confounders.

### 4. Corrections for medication use - Add-back correction

Several cardiometabolic biomarkers examined in this study are influenced by pharmacological treatment. We therefore applied an add-back correction to approximate untreated biomarker levels among participants using relevant medications. The add-back approach assumes treatment lowers biomarker values by an approximately known average amount and adds this amount back to treated individuals to approximate their underlying untreated levels. This method was formalised by Tobin et al. (2005) and is widely used in observational epidemiology for biomarkers affected by medication (Choi et al., 2022). Medication use at the NCDS age 62 sweep was classified using British National Formulary (BNF) subchapters (Table A8).

The total/HDL cholesterol ratio is influenced primarily by lipid-lowering medications, which reduce total cholesterol substantially but have modest effects on HDL cholesterol. Meta-analyses of statin trials suggest total cholesterol reductions of roughly 17–35% depending on drug and dose (Edwards & Moore, 2003; Law et al., 2003). Following prior observational work, we assumed an average 25% reduction in total cholesterol among statin users. For participants reporting lipid-lowering medication (BNF 0212), untreated total cholesterol was estimated by dividing the observed value by 0.75; HDL cholesterol was not corrected. The total/HDL ratio was then computed using corrected total cholesterol and observed HDL.

HbA1c reflects long-term glycaemic control and is directly affected by glucose-lowering medications. Evidence suggests treatment reduces HbA1c by around 0.8–1.5 percentage points on average and adding 1 percentage point is commonly used as a pragmatic correction (Tobin et al., 2005; Eastwood et al., 2016). Accordingly, for participants reporting diabetes medication (BNF 0601), we added 1 percentage point to observed HbA1c.

NT-proBNP is not a direct treatment target and may be influenced by multiple cardiovascular medication classes with heterogeneous, severity-dependent effects (Ponikowski et al., 2016; Yancy et al., 2017; Januzzi et al., 2018). As no widely accepted correction exists, NT-proBNP was analysed using observed values only.

Distributions of uncorrected and corrected cholesterol ratio and HbA1c are shown in Table A9; corrected values were higher on average, consistent with the downward influence of lipid- and glucose-lowering treatment on measured biomarker levels.

**Table A8.** British National Formulary (BNF) medication groups used for biomarker corrections.

| Biomarker | Medication category | BNF chapter | BNF subchapter | Participants using medication, n (%) |
| --- | --- | --- | --- | --- |
| Cholesterol ratio | Lipid-regulating drugs | Cardiovascular system (CH2) | 0212 | 1,814 (36) |
| *Glycated hemoglobin (HbA1c)* | Drugs used in diabetes | Endocrine system (CH6) | 0601 | 561 (11) |
| NT-proBNP | Hypertension and Heart Failure | Cardiovascular system (CH2) | 0205 | 1,463 (30) |
|  | Diuretics | Cardiovascular system (CH2) | 0202 | 372 (8) |
|  | Beta-Adrenoceptor Blocking Drugs | Cardiovascular system (CH2) | 0204 | 571 (12) |

Notes: Medication categories followed the British National Formulary https://www.eclipsesolutions.org/cornwall/bnf.aspx

**Table A9.**
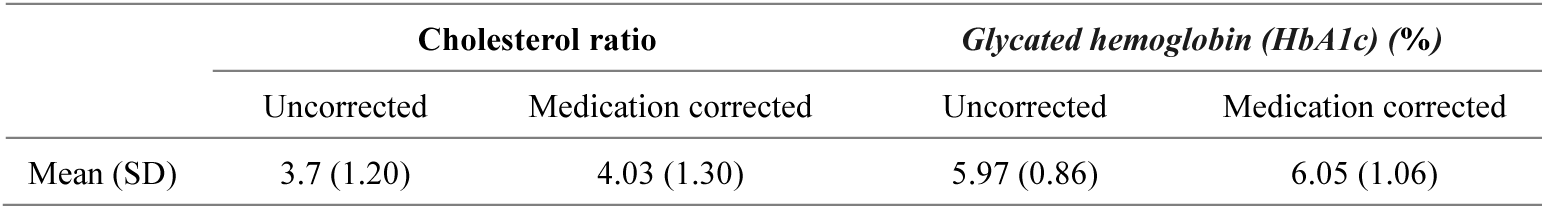
Uncorrected and corrected cholesterol ratio and *Glycated hemoglobin (HbA1c)*

|  | Cholesterol ratio |  | *Glycated hemoglobin (HbA1c) (%)* |  |
| --- | --- | --- | --- | --- |
|  | Uncorrected | Medication corrected | Uncorrected | Medication corrected |
| Mean (SD) | 3.7 (1.20) | 4.03 (1.30) | 5.97 (0.86) | 6.05 (1.06) |

### 5. Sensitivity analyses

#### A. Alternative outcome specifications

To assess whether our conclusions were sensitive to outcome operationalisation, we re-estimated the mediation models using alternative specifications for key biomarkers. CRP was modelled as a continuous outcome (mg/L) and values above 10 mg/L were set to missing to exclude likely acute infection. Cholesterol ratio and HbA1c were dichotomised to indicate elevated risk (cholesterol ratio >6) and diabetes (HbA1c ≥6.5%), respectively. Results were consistent with the main analyses (Tables A10–A12)

**Table A10.**
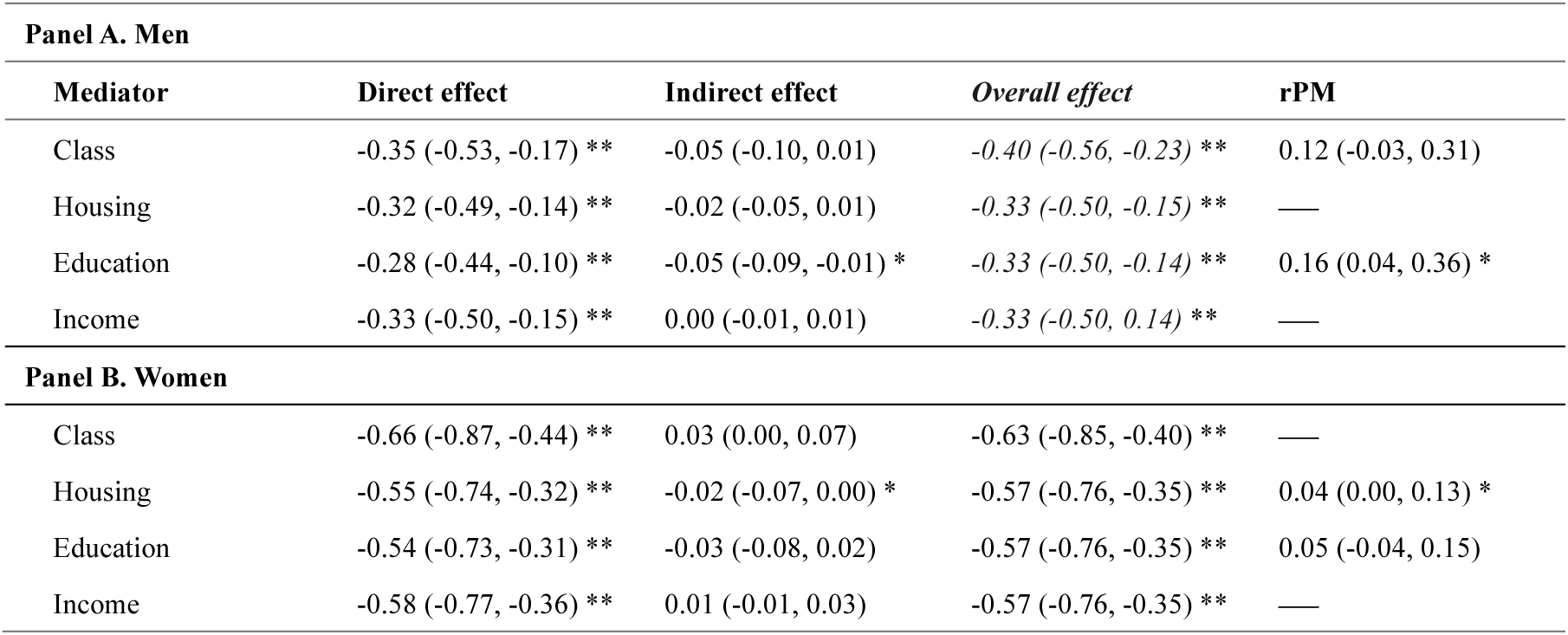
Interventional direct and indirect effects of childhood social class on C-reactive protein at age 62.

**Table A11.**
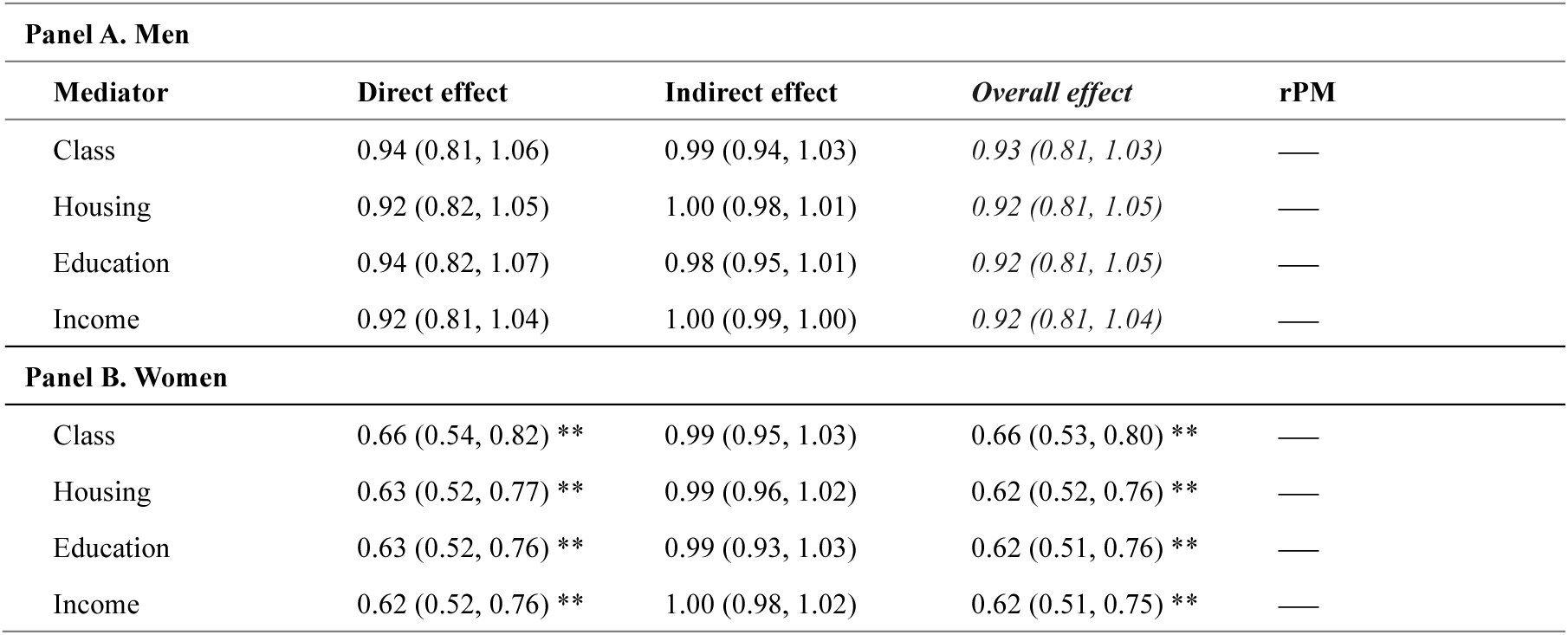
Interventional direct and indirect effects of childhood social class on Cholesterol ratio at age 62.

**Table A12.** Interventional direct and indirect effects of childhood social class on Glycated haemoglobin (HbA1c) at age 62.

| **Panel A. Men** |  |  |  |  |
| --- | --- | --- | --- | --- |
| **Mediator** | **Direct effect** | **Indirect effect** | **Overall effect** | **rPM** |
| Class | 0.78 (0.59, 1.03) | 0.97 (0.89, 1.07) | 0.76 (0.59, 1.00) * | — |
| Housing | 0.76 (0.58, 0.98) * | 0.98 (0.92, 1.01) | 0.74 (0.57, 0.95) * | — |
| Education | 0.77 (0.58, 0.97) * | 0.96 (0.91, 1.03) | 0.74 (0.57, 0.95) * | — |
| Income | 0.74 (0.57, 0.95) * | 1.00 (0.98, 1.01) | 0.74 (0.57, 0.95) * | — |
| **Panel B. Women** |  |  |  |  |
| Class | 0.71 (0.46, 1.11) | 0.94 (0.86, 1.01) | 0.67 (0.42, 1.04) | — |
| Housing | 0.70 (0.44, 0.95) * | 0.94 (0.88, 0.99) * | 0.66 (0.43, 0.89) * | 0.12 (-0.01, 0.44) |
| Education | 0.69 (0.44, 0.97) * | 0.93 (0.87, 1.01) | 0.65 (0.43, 0.88) * | — |
| Income | 0.66 (0.43, 0.89) * | 1.00 (0.97, 1.03) | 0.66 (0.43, 0.90) * | — |

#### B. Alternative medication assumptions and restrictions

To assess sensitivity to medication-related assumptions, we repeated the lipid analyses under alternative statin adjustment factors and conducted restriction-based analyses for NT-proBNP. Specifically, we assumed 20% and 30% reductions in total cholesterol among statin users (instead of 25%) and re-estimated NT-proBNP models excluding participants prescribed heart-failure–related medications. Conclusions were unchanged (Tables A13–A14)

**Table A13.** Interventional direct and indirect effects of childhood social class on cholesterol ratio at age 62, under alternative medication effect.

| 20% reduction in total cholesterol |  |  |  |  |
| --- | --- | --- | --- | --- |
| Panel A. Men |  |  |  |  |
| Mediator | Direct effect | Indirect effect | Overall effect | rPM |
| Class | -0.18 (-0.35, 0.03) | -0.01 (-0.07, 0.05) | -0.19 (-0.35, -0.01) * | — |
| Housing | -0.17 (-0.30, 0.01) | -0.02 (-0.05, 0.00) | -0.19 (-0.32, -0.01) * | — |
| Education | -0.14 (-0.27, 0.03) | -0.05 (-0.09, -0.02) ** | -0.19 (-0.32, -0.01) * | — |
| Income | -0.19 (-0.33, -0.01) * | 0.00 (-0.01, 0.01) | -0.19 (-0.32, -0.01) * | — |
| Panel B. Women |  |  |  |  |
| Class | -0.36 (-0.49, -0.21) ** | 0.00 (-0.03, 0.02) | -0.37 (-0.49, -0.22) ** | — |
| Housing | -0.38 (-0.49, -0.23) ** | -0.01 (-0.03, 0.00) | -0.39 (-0.49, -0.26) ** | — |
| Education | -0.39 (-0.51, -0.25) *** | 0.00 (-0.03, 0.03) | -0.39 (-0.50, -0.26) *** | — |
| Income | -0.39 (-0.50, -0.25) ** | 0.00 (-0.01, 0.01) | -0.39 (-0.49, -0.25) ** | — |
| 30% reduction in total cholesterol |  |  |  |  |
| Panel A. Men |  |  |  |  |
| Class | -0.21 (-0.42, 0.04) | -0.01 (-0.08, 0.05) | -0.22 (-0.41, -0.02) * | — |
| Housing | -0.20 (-0.36, 0.00) * | -0.02 (-0.05, 0.01) | -0.22 (-0.38, -0.01) * | — |
| Education | -0.17 (-0.34, 0.03) | -0.05 (-0.10, -0.02) ** | -0.22 (-0.38, -0.01) * | 0.25 (0.03, 1.80) * |
| Income | -0.22 (-0.39, -0.01) * | 0.00 (-0.01, 0.01) | -0.22 (-0.38, -0.01) * | — |
| Panel B. Women |  |  |  |  |
| Class | -0.42 (-0.57, -0.26) ** | -0.01 (-0.03, 0.02) | -0.43 (-0.58, -0.26) ** | — |
| Housing | -0.45 (-0.57, -0.29) ** | -0.01 (-0.03, 0.00) | -0.46 (-0.58, -0.31) ** | — |
| Education | -0.46 (-0.59, -0.31) ** | 0.00 (-0.03, 0.03) | -0.46 (-0.59, -0.31) ** | — |
| Income | -0.46 (-0.58, -0.31) ** | 0.00 (-0.01, 0.01) | -0.46 (-0.58, -0.31) ** | — |

**Table A14.**
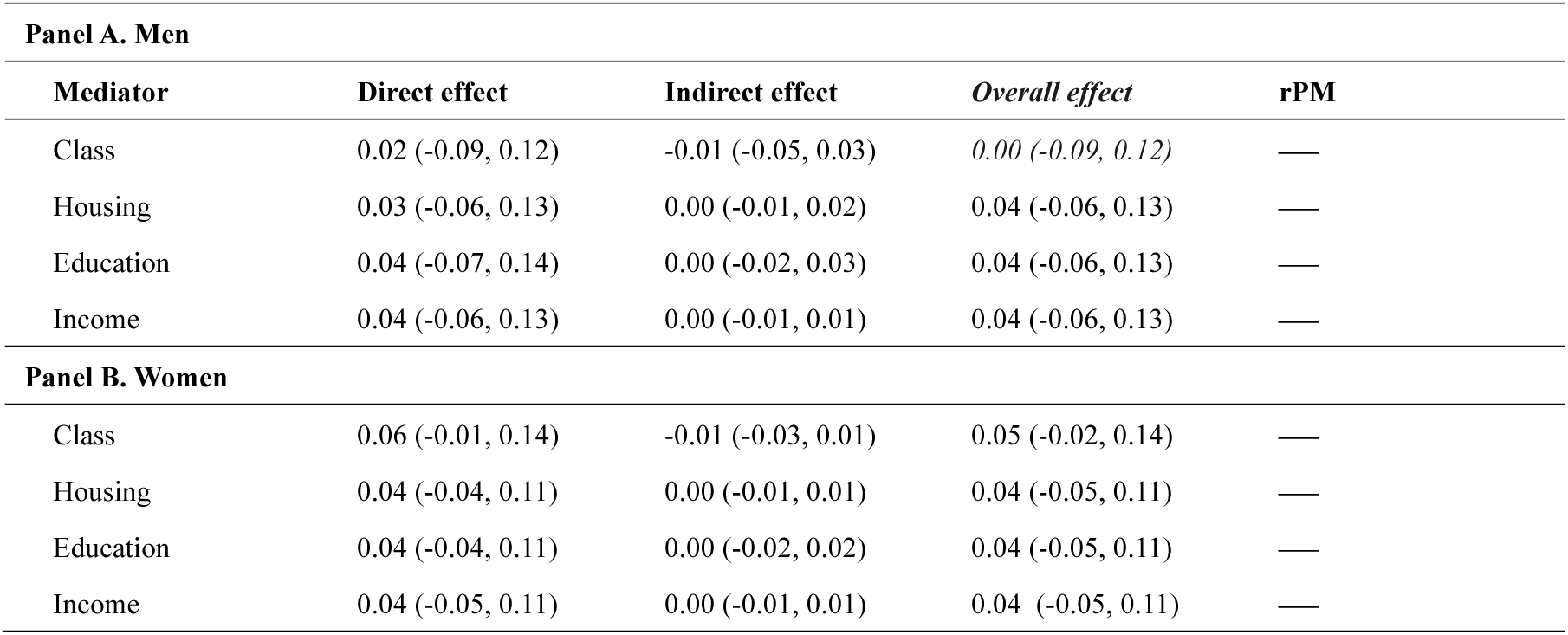
Interventional direct and indirect effects of childhood social class on NT-proBNP at age 62, excluding participants prescribed heart-failure–related medications.

| **Panel A. Men** |  |  |  |  |
| --- | --- | --- | --- | --- |
| **Mediator** | **Direct effect** | **Indirect effect** | **Overall effect** | **rPM** |
| Class | 0.02 (-0.09, 0.12) | -0.01 (-0.05, 0.03) | 0.00 (-0.09, 0.12) | — |
| Housing | 0.03 (-0.06, 0.13) | 0.00 (-0.01, 0.02) | 0.04 (-0.06, 0.13) | — |
| Education | 0.04 (-0.07, 0.14) | 0.00 (-0.02, 0.03) | 0.04 (-0.06, 0.13) | — |
| Income | 0.04 (-0.06, 0.13) | 0.00 (-0.01, 0.01) | 0.04 (-0.06, 0.13) | — |
| **Panel B. Women** |  |  |  |  |
| Class | 0.06 (-0.01, 0.14) | -0.01 (-0.03, 0.01) | 0.05 (-0.02, 0.14) | — |
| Housing | 0.04 (-0.04, 0.11) | 0.00 (-0.01, 0.01) | 0.04 (-0.05, 0.11) | — |
| Education | 0.04 (-0.04, 0.11) | 0.00 (-0.02, 0.02) | 0.04 (-0.05, 0.11) | — |
| Income | 0.04 (-0.05, 0.11) | 0.00 (-0.01, 0.01) | 0.04 (-0.05, 0.11) | — |

#### C. Sample restrictions

To assess sensitivity to sample composition and timing of biomarker measurement, we conducted two restriction-based analyses (Tables A15–A17). First, because COVID-19 disrupted fieldwork and introduced variation in age at assessment, we repeated biomarker models restricting to participants whose biomarkers were collected in 2022–2023 (Table A15–A16; see table notes for analytic sample sizes).

Second, to assess potential selection into biomedical participation, we re-estimated the self-rated health models restricting the sample to respondents who also provided a blood sample, aligning the SRH analytic sample with the biomarker sample (Table A17; see table notes for analytic sample sizes). Estimates were highly comparable to those from the full SRH sample; some direct effect estimates among men were slightly attenuated and confidence intervals widened, consistent with reduced sample size, but the overall mediation pattern and indirect effects were substantively unchanged

**Table A15.** Interventional direct and indirect effects of childhood social class on *Glycated hemoglobin (HbA1c)* at age 62, restricted to biomarker assessments in 2022–2023.

| **Panel A. Men** |  |  |  |  |
| --- | --- | --- | --- | --- |
| **Mediator** | **Direct effect** | **Indirect effect** | **Overall effect** | **rPM** |
| Class | -0.14 (-0.27, 0.02) | -0.02 (-0.06, 0.02) | -0.16 (-0.28, -0.03) * | — |
| Housing | -0.14 (-0.26, -0.01) * | -0.01 (-0.03, 0.01) | -0.15 (-0.27, -0.03) * | — |
| Education | -0.14 (-0.26, -0.01) * | -0.01 (-0.03, 0.01) | -0.15 (-0.27, -0.03) * | — |
| Income | -0.15 (-0.27, -0.02) * | 0.00 (-0.01, 0.01) | -0.15 (-0.27, -0.03) * | — |
| **Panel B. Women** |  |  |  |  |
| Class | -0.13 (-0.21, -0.06) ** | -0.01 (-0.02, 0.00) | -0.13 (-0.21, -0.07) ** | — |
| Housing | -0.10 (-0.17, -0.03) * | -0.01 (-0.01, 0.00) | -0.10 (-0.18, -0.04) ** | — |
| Education | -0.1 (-0.18, -0.03) ** | 0.0 (-0.02, 0.02) | -0.1 (-0.17, -0.04) ** | — |
| Income | -0.10 (-0.17, -0.04) ** | 0.00 (-0.01, 0.00) | -0.10 (-0.17, -0.04) ** | — |

Notes: Analytic sample sizes were 2,041 men and 2,075 women. Models treating adult social class as the mediator excluded participants who reported being out of the labour force at age 55, resulting in smaller samples of 1,827 men and 1,694 women.

**Table A16.** Interventional direct and indirect effects of childhood social class on Cholesterol ratio at age 62, restricted to biomarker assessments in 2022–2023.

| **Panel A. Men** |  |  |  |  |
| --- | --- | --- | --- | --- |
| **Mediator** | **Direct effect** | **Indirect effect** | **Overall effect** | **rPM** |
| Class | -0.16 (-0.33, -0.04) ** | 0.00 (-0.04, 0.06) | -0.15 (-0.31, -0.05) * | — |
| Housing | -0.15 (-0.26, -0.02) * | -0.01 (-0.04, 0.01) | -0.16 (-0.27, -0.04) * | — |
| Education | -0.13 (-0.25, 0.01) | -0.03 (-0.06, -0.01) ** | -0.16 (-0.27, -0.03) * | — |
| Income | -0.16 (-0.27, -0.03) * | 0.00 (-0.01, 0.01) | -0.16 (-0.27, -0.03) * | — |
| **Panel B. Women** |  |  |  |  |
| Class | -0.31 (-0.43, -0.19) ** | 0.00 (-0.03, 0.02) | -0.32 (-0.43, -0.19) ** | — |
| Housing | -0.30 (-0.41, -0.18) ** | -0.01 (-0.03, 0.00) | -0.31 (-0.41, -0.19) ** | — |
| Education | -0.32 (-0.43, -0.20) ** | 0.01 (-0.02, 0.04) | -0.31 (-0.42, -0.19) ** | — |
| Income | -0.31 (-0.42, -0.19) ** | 0.00 (-0.01, 0.01) | -0.31 (-0.42, -0.19) ** | — |

Notes: Analytic sample sizes were 2,055 men and 2,095 women. In models using adult social class as the mediator, exclusion of participants out of the labour force at age 55 reduced the analytic samples to 1,840 men and 1,710 women.

**Table A17.** Interventional direct and indirect effects of childhood social class on self-rated health at age 62, restricted to blood-sample providers.

| **Panel A. Men** |  |  |  |  |
| --- | --- | --- | --- | --- |
| **Mediator** | **Direct effect** | **Indirect effect** | **Overall effect** | **rPM** |
| Class | 0.92 (0.78, 1.09) | 0.87 (0.82, 0.93) ** | 0.81 (0.69, 0.95) * | 0.63 (0.22, 3.73) * |
| Housing | 0.81 (0.68, 0.94) * | 0.96 (0.92, 1.00) | 0.77 (0.65, 0.92) ** | 0.16 (-0.01, 0.41) |
| Education | 0.83 (0.69, 0.98) * | 0.93 (0.89, 0.97) ** | 0.77 (0.65, 0.92) ** | 0.27 (0.09, 0.83) ** |
| Income | 0.78 (0.66, 0.93) ** | 0.99 (0.97, 1.00) | 0.77 (0.65, 0.92) ** | 0.04 (-0.01, 0.15) |
| **Panel B. Women** |  |  |  |  |
| Class | 0.67 (0.52, 0.81) ** | 0.95 (0.90, 0.99) * | 0.63 (0.50, 0.78) ** | 0.09 (0.02, 0.26) * |
| Housing | 0.65 (0.55, 0.77) ** | 0.95 (0.92, 0.99) * | 0.62 (0.52, 0.74) ** | 0.08 (0.02, 0.17) * |
| Education | 0.65 (0.54, 0.77) ** | 0.96 (0.92, 1.00) * | 0.62 (0.52, 0.74) ** | 0.08 (0.01, 0.18) * |
| Income | 0.63 (0.53, 0.75) ** | 0.98 (0.95, 1.00) * | 0.62 (0.52, 0.74) ** | 0.03 (0.00, 0.09) * |

Notes: Analytic sample sizes for self-rated health at age 62 (restricted to blood-sample providers) were 3,146 men and 3,160 women. In models using adult social class as the mediator, participants reporting being out of the labour force at age 55 were excluded, reducing the analytic samples to 2,791 men and 2,550 women.

#### D. Income mediation robustness checks

Because mediation via income appeared weaker than for other adult SEP indicators, we assessed whether this reflected how income was operationalised and measured rather than substantive differences in its mediating role. First, to place income on contrasts more comparable to the binary mediators and to allow for potential non-linearity, we re-estimated models using three pre-specified binary operationalisations of household income (relative advantage, low income, and a poverty-proxy threshold; Tables A18–A19). Second, to assess sensitivity to income measurement at age 55, we repeated analyses using household income measured at age 50 (Table A20-A21).

Income mediation remained limited in sensitivity analyses. Replacing continuous log-income with binary income measures yielded little change in indirect effects for most outcomes; the only notable difference was a small increase in mediation for men’s self-rated health when higher income was operationalised as belonging to the top 40% (IIE RR = 0.97; rPM = 0.09) (Table A18). Using household income measured at age 50 produced a similarly modest mediated component for men’s self-rated health (IIE RR = 0.98; rPM = 0.07), while conclusions for women and for biomarkers were unchanged (Table A20-A21)

**Table A18.** Interventional direct and indirect effects of childhood social class on health outcomes age 62 among men, using binary household income as the mediator.

| Men | Direct effect | Indirect effect | Overall effect | rPM |
| --- | --- | --- | --- | --- |
| **Self-rated health RR (CI)** |  |  |  |  |
| Log-income age 55 | 0.78 (0.68, 0.88) ** | 0.99 (0.97, 1.00) | 0.77 (0.67, 0.88) ** | 0.04 (0.00, 0.11) |
| high = top 40% | 0.79 (0.69, 0.90) *** | 0.97 (0.95, 0.99) *** | 0.77 (0.66, 0.87) *** | 0.09 (0.04, 0.23) *** |
| high = top 80% | 0.77 (0.67, 0.88) ** | 1.00 (0.98, 1.01) | 0.77 (0.66, 0.87) ** | — |
| high = $\geq 60\%$ of median | 0.77 (0.68, 0.89) ** | 0.99 (0.98, 1.01) | 0.77 (0.66, 0.87) ** | — |
| **C-reactive protein RR (CI)** |  |  |  |  |
| Log-income age 55 | 0.62 (0.49, 0.85) ** | 1.00 (0.99, 1.01) | 0.62 (0.49, 0.84) ** | — |
| high = top 40% | 0.64 (0.50, 0.82) ** | 0.98 (0.95, 1.01) | 0.63 (0.48, 0.80) ** | 0.03 (-0.02, 0.10) |
| high = top 80% | 0.63 (0.48, 0.80) ** | 1.00 (0.99, 1.04) | 0.63 (0.48, 0.80) ** | — |
| high = $\geq 60\%$ of median | 0.63 (0.49, 0.81) ** | 1.00 (0.98, 1.03) | 0.63 (0.48, 0.80) ** | — |
| **Cholesterol ratio estimates (CI)** |  |  |  |  |
| Log-income age 55 | -0.14 (-0.28, 0.01) | 0.00 (-0.01, 0.01) | -0.14 (-0.28, 0.01) | — |
| high = top 40% | -0.13 (-0.26, 0.00) * | -0.01 (-0.02, 0.00) | -0.14 (-0.27, 0.00) * | — |
| high = top 80% | -0.14 (-0.26, 0.00) * | 0.00 (-0.01, 0.02) | -0.14 (-0.27, 0.00) * | — |
| high = $\geq 60\%$ of median | -0.14 (-0.26, 0.00) * | 0.00 (-0.01, 0.01) | -0.14 (-0.27, 0.00) * | — |
| **Glycated hemoglobin estimates (CI)** |  |  |  |  |
| Log-income age 55 | -0.14 (-0.24, -0.04) * | 0.00 (-0.01, 0.01) | -0.14 (-0.24, -0.03) * | — |
| high = top 40% | -0.14 (-0.22, -0.04) * | 0.00 (-0.01, 0.01) | -0.14 (-0.22, -0.04) ** | — |
| high = top 80% | -0.14 (-0.22, -0.04) * | 0.00 (-0.01, 0.01) | -0.14 (-0.22, -0.04) ** | — |
| high = $\geq 60\%$ of median | -0.14 (-0.22, -0.04) * | 0.00 (-0.01, 0.01) | -0.14 (-0.22, -0.04) ** | — |
| **NT-proBNP estimates (CI)** |  |  |  |  |
| Log-income age 55 | 0.04 (-0.03, 0.14) | 0.00 (-0.01, 0.00) | 0.04 (-0.03, 0.14) | — |
| high = top 40% | 0.05 (-0.03, 0.15) | 0.00 (-0.01, 0.00) | 0.04 (-0.03, 0.14) | — |
| high = top 80% | 0.04 (-0.03, 0.14) | 0.00 (0.00, 0.00) | 0.04 (-0.03, 0.14) | — |
| high = $\geq 60\%$ of median | 0.04 (-0.03, 0.14) | 0.00 (0.00, 0.00) | 0.04 (-0.03, 0.14) | — |

**Table A19.**
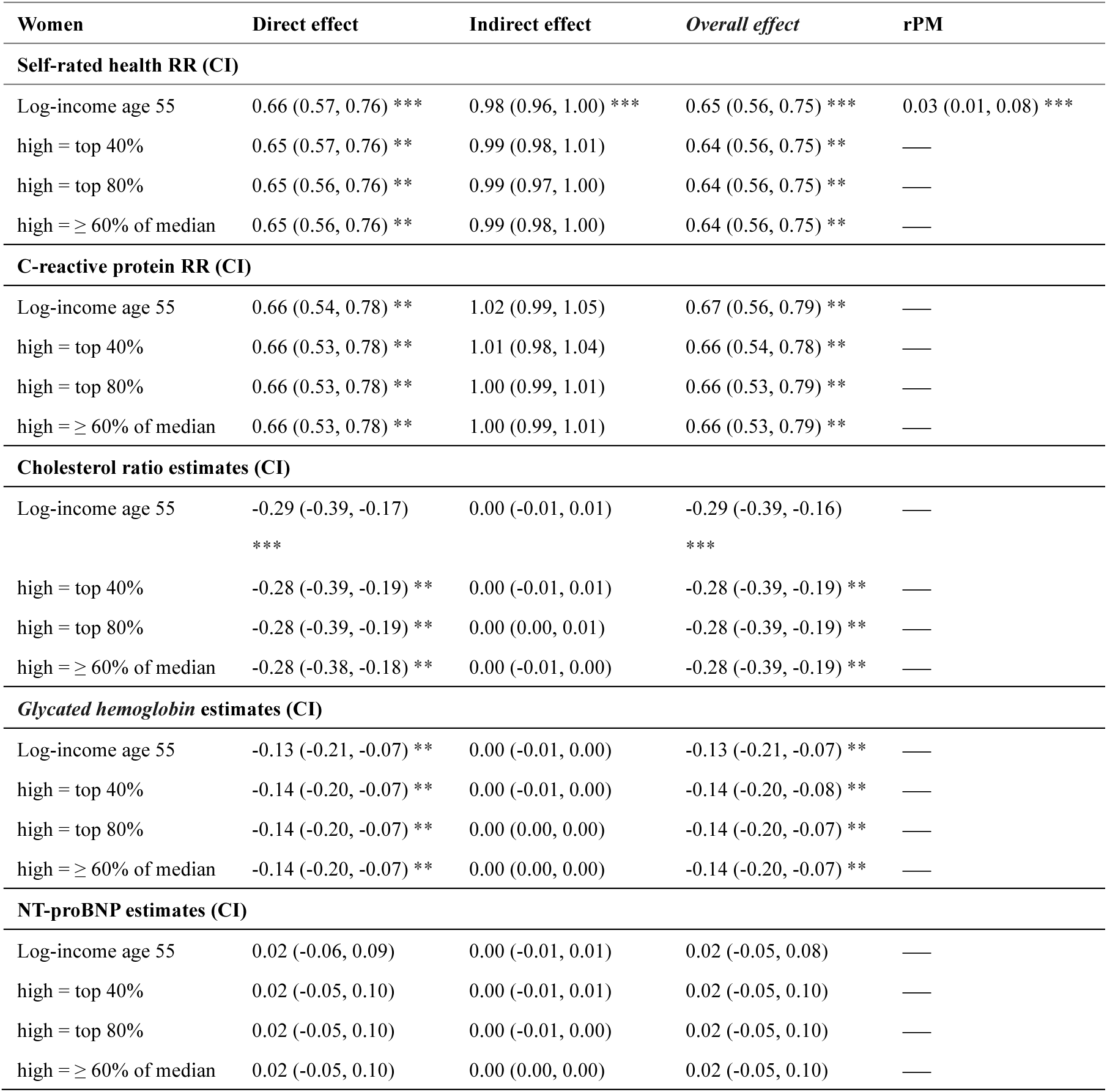
Interventional direct and indirect effects of childhood social class on health outcomes age 62 among women, using binary household income as the mediator.

| **Women** | **Direct effect** | **Indirect effect** | **Overall effect** | **rPM** |
| --- | --- | --- | --- | --- |
| **Self-rated health RR (CI)** |  |  |  |  |
| Log-income age 55 | 0.66 (0.57, 0.76) *** | 0.98 (0.96, 1.00) *** | 0.65 (0.56, 0.75) *** | 0.03 (0.01, 0.08) *** |
| high = top 40% | 0.65 (0.57, 0.76) ** | 0.99 (0.98, 1.01) | 0.64 (0.56, 0.75) ** | — |
| high = top 80% | 0.65 (0.56, 0.76) ** | 0.99 (0.97, 1.00) | 0.64 (0.56, 0.75) ** | — |
| high = ≥ 60% of median | 0.65 (0.56, 0.76) ** | 0.99 (0.98, 1.00) | 0.64 (0.56, 0.75) ** | — |
| **C-reactive protein RR (CI)** |  |  |  |  |
| Log-income age 55 | 0.66 (0.54, 0.78) ** | 1.02 (0.99, 1.05) | 0.67 (0.56, 0.79) ** | — |
| high = top 40% | 0.66 (0.53, 0.78) ** | 1.01 (0.98, 1.04) | 0.66 (0.54, 0.78) ** | — |
| high = top 80% | 0.66 (0.53, 0.78) ** | 1.00 (0.99, 1.01) | 0.66 (0.53, 0.79) ** | — |
| high = ≥ 60% of median | 0.66 (0.53, 0.78) ** | 1.00 (0.99, 1.01) | 0.66 (0.53, 0.79) ** | — |
| **Cholesterol ratio estimates (CI)** |  |  |  |  |
| Log-income age 55 | -0.29 (-0.39, -0.17) *** | 0.00 (-0.01, 0.01) | -0.29 (-0.39, -0.16) *** | — |
| high = top 40% | -0.28 (-0.39, -0.19) ** | 0.00 (-0.01, 0.01) | -0.28 (-0.39, -0.19) ** | — |
| high = top 80% | -0.28 (-0.39, -0.19) ** | 0.00 (0.00, 0.01) | -0.28 (-0.39, -0.19) ** | — |
| high = ≥ 60% of median | -0.28 (-0.38, -0.18) ** | 0.00 (-0.01, 0.00) | -0.28 (-0.39, -0.19) ** | — |
| **Glycated hemoglobin estimates (CI)** |  |  |  |  |
| Log-income age 55 | -0.13 (-0.21, -0.07) ** | 0.00 (-0.01, 0.00) | -0.13 (-0.21, -0.07) ** | — |
| high = top 40% | -0.14 (-0.20, -0.07) ** | 0.00 (-0.01, 0.00) | -0.14 (-0.20, -0.08) ** | — |
| high = top 80% | -0.14 (-0.20, -0.07) ** | 0.00 (0.00, 0.00) | -0.14 (-0.20, -0.07) ** | — |
| high = ≥ 60% of median | -0.14 (-0.20, -0.07) ** | 0.00 (0.00, 0.00) | -0.14 (-0.20, -0.07) ** | — |
| **NT-proBNP estimates (CI)** |  |  |  |  |
| Log-income age 55 | 0.02 (-0.06, 0.09) | 0.00 (-0.01, 0.01) | 0.02 (-0.05, 0.08) | — |
| high = top 40% | 0.02 (-0.05, 0.10) | 0.00 (-0.01, 0.01) | 0.02 (-0.05, 0.10) | — |
| high = top 80% | 0.02 (-0.05, 0.10) | 0.00 (-0.01, 0.00) | 0.02 (-0.05, 0.10) | — |
| high = ≥ 60% of median | 0.02 (-0.05, 0.10) | 0.00 (0.00, 0.00) | 0.02 (-0.05, 0.10) | — |

**Table A20.**
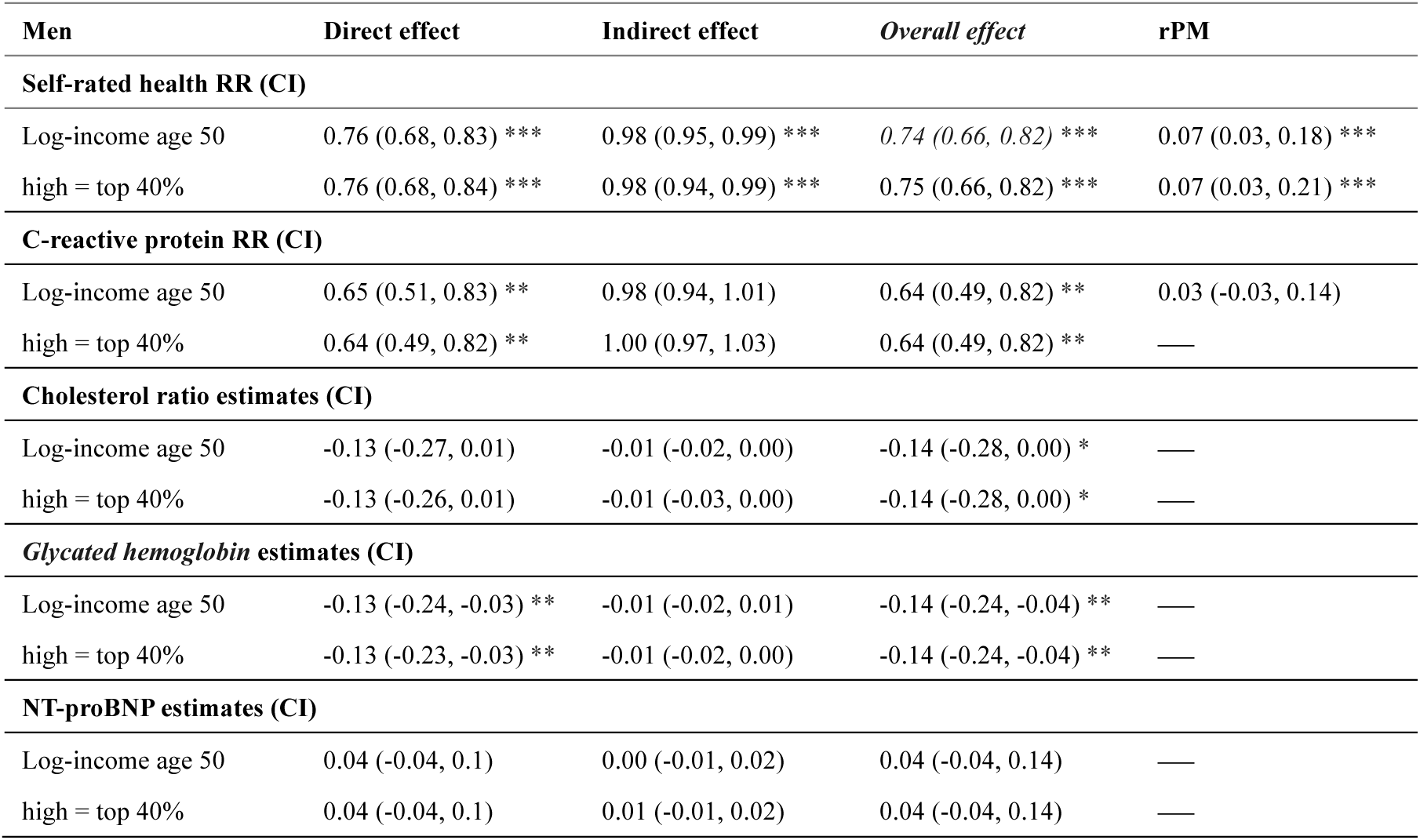
Interventional direct and indirect effects of childhood social class on health outcomes age 62 among men, using household income at age 50 as the mediator.

| Men | Direct effect | Indirect effect | Overall effect | rPM |
| --- | --- | --- | --- | --- |
| **Self-rated health RR (CI)** |  |  |  |  |
| Log-income age 50 | 0.76 (0.68, 0.83) *** | 0.98 (0.95, 0.99) *** | 0.74 (0.66, 0.82) *** | 0.07 (0.03, 0.18) *** |
| high = top 40% | 0.76 (0.68, 0.84) *** | 0.98 (0.94, 0.99) *** | 0.75 (0.66, 0.82) *** | 0.07 (0.03, 0.21) *** |
| **C-reactive protein RR (CI)** |  |  |  |  |
| Log-income age 50 | 0.65 (0.51, 0.83) ** | 0.98 (0.94, 1.01) | 0.64 (0.49, 0.82) ** | 0.03 (-0.03, 0.14) |
| high = top 40% | 0.64 (0.49, 0.82) ** | 1.00 (0.97, 1.03) | 0.64 (0.49, 0.82) ** | — |
| **Cholesterol ratio estimates (CI)** |  |  |  |  |
| Log-income age 50 | -0.13 (-0.27, 0.01) | -0.01 (-0.02, 0.00) | -0.14 (-0.28, 0.00) * | — |
| high = top 40% | -0.13 (-0.26, 0.01) | -0.01 (-0.03, 0.00) | -0.14 (-0.28, 0.00) * | — |
| **Glycated hemoglobin estimates (CI)** |  |  |  |  |
| Log-income age 50 | -0.13 (-0.24, -0.03) ** | -0.01 (-0.02, 0.01) | -0.14 (-0.24, -0.04) ** | — |
| high = top 40% | -0.13 (-0.23, -0.03) ** | -0.01 (-0.02, 0.00) | -0.14 (-0.24, -0.04) ** | — |
| **NT-proBNP estimates (CI)** |  |  |  |  |
| Log-income age 50 | 0.04 (-0.04, 0.1) | 0.00 (-0.01, 0.02) | 0.04 (-0.04, 0.14) | — |
| high = top 40% | 0.04 (-0.04, 0.1) | 0.01 (-0.01, 0.02) | 0.04 (-0.04, 0.14) | — |

**Table A21.** Interventional direct and indirect effects of childhood social class on health outcomes age 62 among women, using household income at age 50 as the mediator.

| Women | Direct effect | Indirect effect | Overall effect | rPM |
| --- | --- | --- | --- | --- |
| **Self-rated health RR (CI)** |  |  |  |  |
| Log-income age 50 | 0.65 (0.56, 0.75) ** | 1.00 (0.99, 1.00) | 0.65 (0.56, 0.74) ** | — |
| high = top 40% | 0.65 (0.56, 0.75) ** | 0.99 (0.98, 1.00) | 0.65 (0.56, 0.74) ** | — |
| **C-reactive protein RR (CI)** |  |  |  |  |
| Log-income age 50 | 0.63 (0.54, 0.73) ** | 1.00 (0.99, 1.01) | 0.63 (0.54, 0.72) ** | — |
| high = top 40% | 0.66 (0.54, 0.81) ** | 1.00 (0.98, 1.01) | 0.66 (0.54, 0.81) ** | — |
| **Cholesterol ratio estimates (CI)** |  |  |  |  |
| Log-income age 50 | -0.29 (-0.38, -0.18) ** | 0.00 (-0.01, 0.00) | -0.29 (-0.39, -0.18) ** | — |
| high = top 40% | -0.29 (-0.39, -0.19) ** | 0.00 (-0.01, 0.00) | -0.29 (-0.39, -0.18) ** | — |
| **Glycated hemoglobin estimates (CI)** |  |  |  |  |
| Log-income age 50 | -0.13 (-0.21, -0.07) ** | 0.00 (-0.01, 0.00) | -0.14 (-0.21, -0.07) ** | — |
| high = top 40% | -0.13 (-0.21, -0.07) ** | 0.00 (-0.01, 0.00) | -0.14 (-0.21, -0.07) ** | — |
| **NT-proBNP estimates (CI)** |  |  |  |  |
| Log-income age 50 | 0.02 (-0.05, 0.08) | 0.00 (0.00, 0.00) | 0.02 (-0.05, 0.09) | — |
| high = top 40% | 0.02 (-0.05, 0.09) | 0.00 (-0.01, 0.00) | 0.02 (-0.05, 0.09) | — |

